# Mass Testing with Contact Tracing Compared to Test and Trace for Effective Suppression of COVID-19 in the UK: A rapid review

**DOI:** 10.1101/2021.01.13.21249749

**Authors:** Mathew Mbwogge

## Abstract

**Background:** Making testing available to everyone and tracing contacts might be the gold standard towards the control of COVID-19, particularly when significant transmissions are without symptoms. This study evaluated the effectiveness of mass testing and contact tracing in the suppression of COVID-19 compared to conventional Test and Trace in the UK.

**Design:** A rapid review of available evidence

**Primary research question:** Is there evidence that mass testing and tracing could suppress community spread of SARS-CoV-2 infections better than Test and Trace?

**Secondary research question:** What is the proportion of asymptomatic cases of SARS-CoV-2 reported during mass testing interventions?

**Methods:** Literature was searched in September through December 2020 in Google Scholar, ScienceDirect, Mendeley and PubMed.

**Results:** Literature search yielded 286 articles from Google Scholar, 20 from Science Direct, 14 from Mendeley, 27 from Pubmed and 15 through manual search. Altogether 35 articles were included, making a sample size of close to a million participants.

**Conclusion:** There was a very low level but promising evidence of 76.9% (95% CI: 46.2 – 95.0, P=0.09) majority vote in favour of the intervention under the primary objective. The overall proportion of asymptomatic cases among those tested positive and tested sample populations under the secondary objective was 40.7% (95% CI: 38.8– 42.5) and 0.01% (95% CI: 0.01 – 0.012) respectively. Conventional test and trace should be superseded by a decentralised and regular mass rapid testing and contact tracing, championed by GP surgeries and low cost community services.

## Introduction

UK’s Test and Trace has been suboptimal in addressing the testing needs of those infected with SARS-CoV-2, let alone handling its new variant(1). The panic over rising cases and a potentially more dangerous second wave led to the creation of the National Institute for Health Protection(2). Follow-up measures have been national lockdown, increased testing, tier system, furlough scheme and approval of the Pfizer, Oxford AstraZenaca and Moderna vaccines(3, 4). As part of the above, about 56 million tests have been performed as at January 10, 2021, with about 1.3 million vaccinated(5). The plans to launch the 100 billion pound “moonshot” programme will only sound as good if tests are delivered based on infections rather than on symptoms(6, 7). I concur with the Director General of WHO that “*you cannot fight a fire blindfolded. And we cannot stop this pandemic if we don’t know who is infected*”(8). Infections could better inform public policy and facilitate equitable rollout of vaccines. While hoping that vaccines will proof as effective as deemed in the development of herd immunity, it is important not to lose sight of other control measures. Regular mass testing combined with contact tracing could be a novel control strategy not just to inform vaccination but also to guard against uncertainties arising from any new variant(9).

## Research in Context

**Prior to this study**

Three modelling studies implemented in the UK dealing with mass testing were found. One of the models did a feasibility analysis of mass testing as a lockdown exit strategy. One model compared mass testing with symptom-based test and trace, while another model compared mass testing with symptom-based testing and isolation. There have been no real-time study in the UK comparing mass test and trace with the conventional test and trace system, reason being that mass testing and contact tracing was judged to be impossible. One systematic review was found, evaluating the effectiveness of universal screening for SARS-CoV-2 compared to no screening.

**In this study**

This is the first review to the best of my knowledge that sought to evaluate the benefits of mass testing and contact tracing (hybrid strategy) compared to test and trace, in the control of COVID-19 in the UK. The proportion of asymptomatic cases has also been explored.

**Way forward**

There is urgent need for a strategy that will identify SARS-CoV-2 carriers when their viral load is high and are most likely to be infectious. Real-time studies are needed to (1) get a true picture of disease burden, (2) validate various mass testing options and (3) better inform vaccination programme and other control measures.

## Definitions

**Contact Tracing:** The process of reporting, identifying, listing and follow-up of individuals that have come in contact with those told to self-isolation due to positive COVID-19 test results, in view of asking them to quarantine(10–12).

**A Contact:** A COVID-19 contact is defined as any person who has been in close contact with a COVID-19 confirmed case either 2 days before symptom onset or before sample collection and 14 days after the onset of symptoms or after the sample was taken(10).

**Isolation:** The act of being restrained from social interaction for a period of 10 days from symptom onset, as a result of either exhibiting COVID-19 symptoms or confirmed positive test result(13).

**Quarantine:** To restrict the movement of contacts or people suspected to have come in contact with COVID-19 cases for a period of 14 days from the time of last contact. This includes contacts as defined herein, household members, passengers of inbound flights from high-risk zones(13).

**Test and Trace:** The process of 1) testing COVID-19 symptomatic individuals in view of asking them to stay in isolation and declare contacts, if with positive test results and 2)identifying, listing and follow-up of reported contacts so as to ask them to quarantine(10, 11). It is called Test and Protect in Scotland and Test, Trace and Protect in Wales and Northern Ireland(14–16).

**Mass testing:** Also known as active case finding is the proactive approach of testing individuals irrespective of symptoms so as to track the contacts of those infected and break the transmission circuit of the virus early enough(17).

**Effectiveness:** The extent to which the rolling out of the intervention and/or control benefits the public, economy and environment in terms of limiting the spread of SARS-CoV-2/COVID-19, measured as the reduction in reproduction number-R(18).

**Suppression:** Implementation of a package of control measures in the middle of differential community transmissions of SARS-CoV-2/COVID-19 in view of reasonably bringing the reproduction number (R) below 1, so as to safely reopen the economy(19).

## Conventional Test and Trace (TT) System

Figure 1 below shows the traditional “Test and Trace” system currently implemented in the UK, with a number of possible implications. Kindly refer to www.gov.uk for further details on how the Test and Trace system works(20). In the face of rising asymptomatic infectivity, the present TT delivery strategy can be categorised as “the cake not worth the candle”, since the programme fails to determine the true burden of the disease.

**Figure 1:**
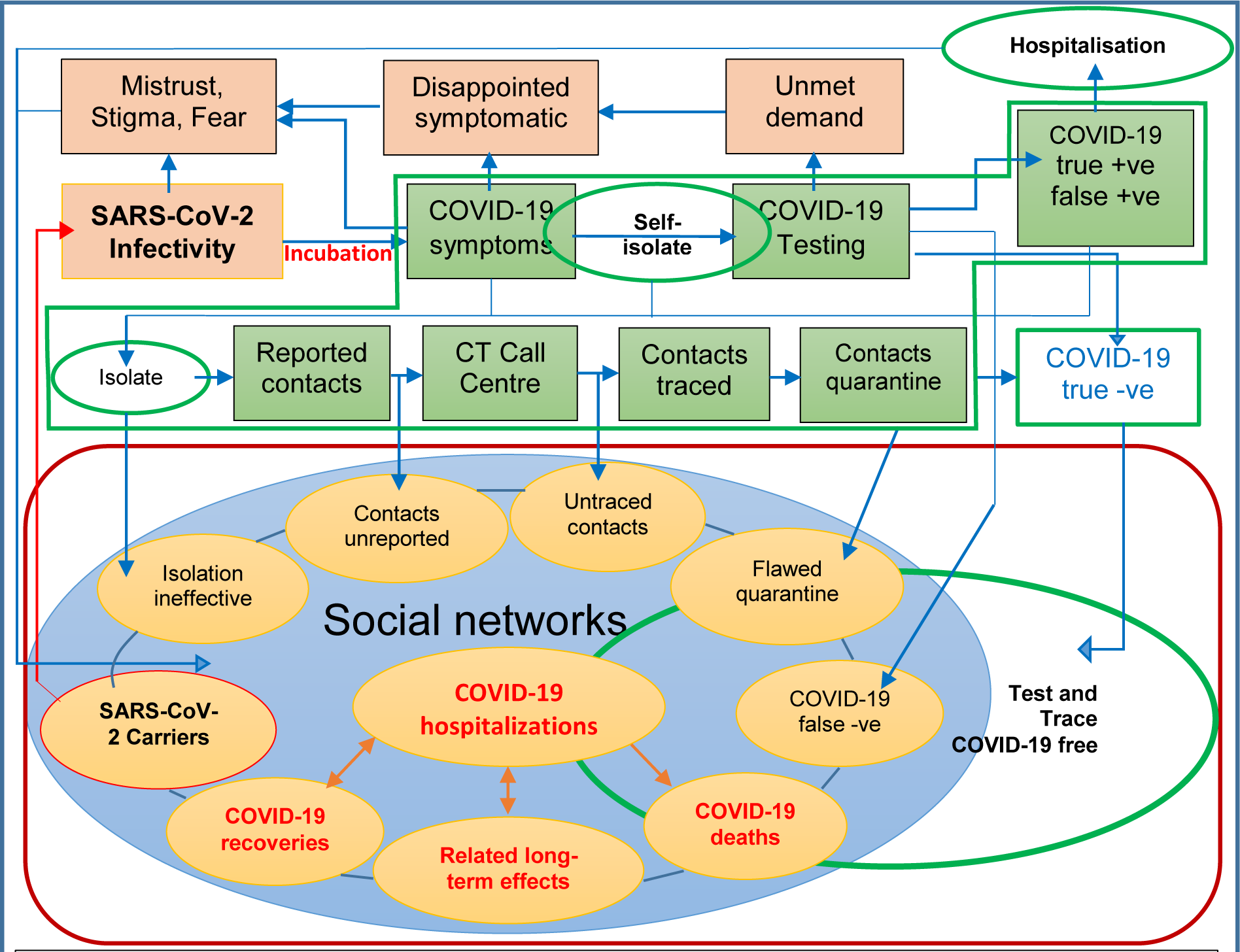
Conventional Test and Trace system: Green panel shows workflow of Test and Trace system SARS-CoV-2: Severe acute respiratory syndrome coronavirus 2 COVID-19: Coronavirus disease 2019; CT: Contact Tracing; +ve: Positive; -ve: Negative

The following can generally be observed from the above conventional system;

1. Individuals who are asymptomatic and presymptomatic are missed out(21–23)
2. People are generally afraid of quarantine and may shy away from TT(24).
3. The decision of public safety about getting tested has been shifted to the public
4. Operational false positive estimates in the UK are currently unknown(25)
5. Proportion of daily asymptomatic cases is still not part of the national statistics
6. Test and trace depends on self-reported contacts which may be flawed
7. A proportion of the public is hesitant due to stigma surrounding data ethics(26)
8. TT is a shift away from Universal Health Coverage, amid a pandemic(27)
9. Long travel among others is a serious barrier to accessing test centres

### The “Infectivity Problem” of COVID-19

The “infectivity problem” can be summarized into 1) Test ramp-up controversy, 2) TT system leakages, 3) Time-to-test paradox and 4) Inequitable test delivery and delays.

**Test Ramp-up Controversy:** This is the heated discussion and lockdown-related antagonism from the public, regarding the undesired positive correlation which was presumed inverse, between testing capacity and COVID-19 cases. The supposed endgame of test ramp-up was to contain the virus but countries have found themselves in the “opposite-of-things”. This may be due to more cases now being detected as a result of increased testing or because the testing is not comprehensive and early enough to outweigh the viral shedding. This may culminate into the UK’s “operation moonshot” controversy if testing rate continues to be less than infectivity rate(28).

**Test and Trace System Leakages:** Leakages refer to the infectious population that apparently was supposed to be tested but end up not being detected. This includes those with either unreported symptoms or not presenting for test, those sent home due to unavailability of tests, asymptomatic and presymptomatic individuals, unreported and untraced contacts, false negatives as well as the non-compliant to isolation and quarantine rules(29, 30).

**Time-to-test Paradox:** This refers to the conflicting interest of whether to test prior to symptoms or upon reported symptoms. The TT programme has been designed not to test people at very early stages of infection for fear of missing out the very cases it is meant to detect. The same is true when people are tested late(31, 32). Research suggest that the serial interval of Covid-19 is shorter than the incubation period, indicating possible infectivity multiplier effect prior to symptom onset(33, 34). This is further compounded by operational false positives and false negatives(25).

**Inequitable Test Delivery and Delays:** This has to do with testing that is not delivered at point-of-care thereby eliminating a certain group of persons, delays in testing those reporting symptoms, test-to-results delay as well as contact tracing time lapse. The aforementioned in addition to not testing those without symptoms led to increasing infections in the face of delivering the highest number of tests in Europe(35). A disease that is as deadly as the present gives no tolerance to turnaround time and mitigation programme mistakes, the biggest of which has been the apparently neglected asymptomatic infectivity.

## How the Intervention Should Work

The novel mass test and contact trace strategy is one that 1) extends the present Test and Trace system to the general public and 2) moving it from laboratory based to point-of-care thereby enhancing acceptability, accessibility and equity. This framework is a modification of that proposed by Lassi et al(36). Community ownership involves each individual registering with a general practitioner (GP) surgery, capacitated to perform routine open invitation testing irrespective of symptoms. The strategy necessitates the availability of rapid easy-to-run cost effective tests and a succinct exit strategy. Inputs include macro policies (fiscal, support schemes, PPE, hygiene and sanitation, environmental, tier system etc.), messa policies (GP capacitation, social gathering, at-risk group, vaccination etc.) and micro policies (health status, personal hygiene, compliance to national guidelines, tracing app acceptability). Routine health checks with GPs have hardly raised concerns around privacy due to trust. It is more reliable and confident for GPs to run testing programmes, offer direct vaccination and therapeutic treatment to those that have tested positive, as well as request those with positive test results to report contacts on the NHS contact tracing platform. Through a shared platform, Contact Tracing Centre (CTC) could be granted access to a limited dataset or transferred to the NHS Contact Tracing system. The CTC liaises with index cases for any additional contacts, and calls all listed contacts for quarantine advice. Based on data collected, the tier management team and environmental health officers work in synergy with local councils towards local containment strategies, similar to how the local outbreak in Leicester was managed. Figure 2 below shows the workflow of proposed framework.

**Figure 2:**
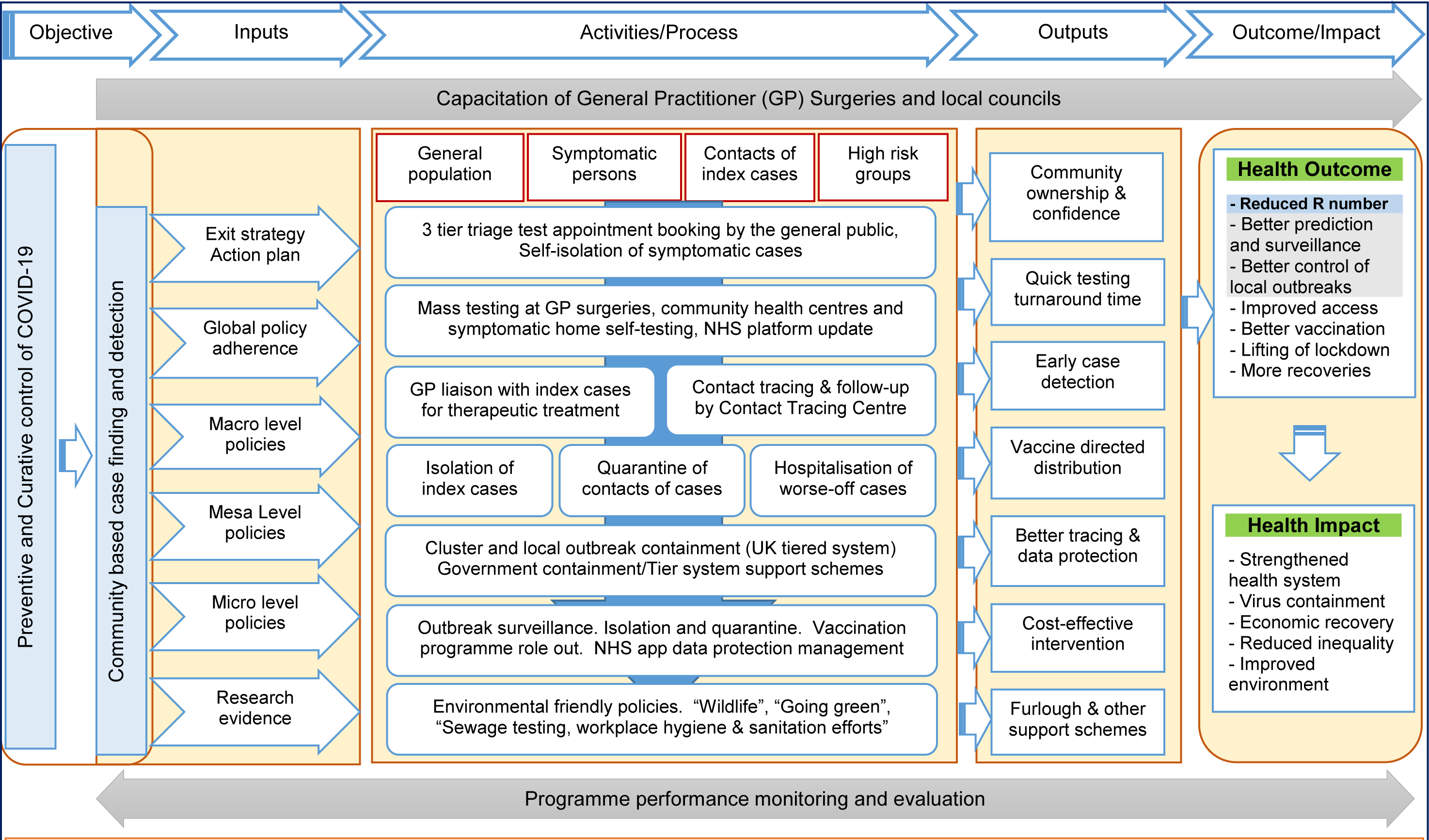
Conceptual framework for decentralised community based mass testing and contact tracing Adapted from Lassi et al (2014): licensed under the Creative Commons Attribution 4.0 International License (http://creativecommons.org/licenses/by/4.0/)

## Objectives

The study objective is twofold. Firstly to evaluate the evidence of Mass Test and Trace compared to Test and Trace in the suppression of community transmissions of SARS-CoV-2/COVID-19. Secondly to find out the proportion of reported asymptomatic carriers during mass testing interventions.

The population of interest included symptomatic and apparently healthy individuals within the UK setting. Coronavirus disease 2019 (COVID-19) is a recently emerged acute respiratory disease caused by a highly pathogenic coronavirus called Severe acute respiratory syndrome coronavirus 2 (SARS-CoV-2) that emerged in 2019 in Wuhan China(37). SARS-CoV-2 shares more than 90% amino acid and 79% genome sequence identities with SARS-CoV and 50% genome identity with the Middle East Respiratory Syndrome coronavirus (MERS-CoV). Symptoms of COVID-19 include prolonged high fever (≥38°C), prolonged cough, dyspnea and loss of taste. The intervention of interest is mass testing irrespective of symptoms and tracing contacts of positive cases, while the control is symptom-based test and trace.

### Primary research question

Is there evidence that testing irrespective of symptoms combined with tracing could suppress SARS-CoV-2 infections better than symptom-based testing and tracing?

### Secondary research question

What is the proportion of asymptomatic carriers of SARS-CoV-2 reported during mass testing interventions?

## Methodology

### Study Outcomes

✓ Effectiveness
✓ Cost-effectiveness
✓ Safety
✓ Acceptability
✓ Equity
✓ Asymptomatic proportion

### Search Strategy

A literature search was performed on September 9, 2020 and constantly refreshed through December 22, 2020. The search involved all English articles published in 2020 including grey literature. Search terms in Google Scholar included “[UK] [Effectiveness of mass testing] [COVID-19] [SARS-CoV-2] [Contact OR tracing] [Contact tracing] [Effectiveness of test and trace] –Animals –Influenza –HIV –Cancer”. An advanced search was performed in ScienceDirect for “[Test and trace] OR [contact tracing] AND [COVID-19] AND [SARS-CoV-2] AND [asymptomatic] AND [symptomatic] OR [screening for SARS-CoV-2] OR [mass testing for SARS-CoV-2]”, whose title included “[UK] AND [test and trace] OR [contact tracing] OR [community screening for SARS-CoV-2] OR [mass testing for SARS-CoV-2]” restricted to the year 2020. A search in PubMed included “((((((((Mass testing for COVID-19 and “Contact tracing”) OR (Mass testing for SARS-CoV-2 and “Contact tracing”)) OR (”Test and trace”)) OR (”Mass testing” and “symptom-based testing”)) NOT (Animals)) NOT (HIV)) NOT (Influenza)) NOT (Ebola)) NOT (Cancer)”. Finally, a search for “Mass testing for COVID-19” AND “contact tracing for COVID-19” OR “mass testing for SARS-CoV-2” AND “contact tracing for SARS-CoV-2” was done in Mendeley.

### Exclusion Criteria

All articles published before the year 2020, non-English articles, articles whose full texts were not accessible, non-COVID-19 articles, articles on non-human subjects and non-mass testing articles. Given that this review was about detecting people currently infected, we excluded antibody studies. We also excluded editorials and protocols

### Eligibility Criteria

Full text articles comparing testing irrespective of symptoms and contact tracing with symptom-based test and trace, as well as any partial comparison between the above.

## Data Management

### Data Extraction

Data extraction was done by a single reviewer who also did a detailed review of extracted data for individual studies. Extracted data included the study date, author, setting, study design, study objective, type of intervention, outcome, type of participants, strategies used, assumptions, data analysis, results, study limitations and bias.

### Criteria for Grouping Studies

In accordance with the study objective and logical framework, studies for synthesis were grouped according to outcome. In order to capture the studies whose interventions geared towards evaluating effects on outcomes of interest(38). This made it easy to articulate synthesis to research questions.

### Standardized and Synthesis Metrics

Direction of effect was used as the standardize metric because there was a lack of precision specific to the effect of intervention and control in the results presented by different studies. This did not permit the calculation of summary statistics(39). In light of the above, vote counting was the best match in synthesising the results. A sign test was used to indicate whether there was an evidence of effect. Equivocal effects between the intervention and control were considered to be distributed around the null hypothesis of no effect. This study made use of Synthesis Without Meta-Analysis (SWiM) reporting guidelines to report review results(40).

### Heterogeneity Assessment

Heterogeneity of studies was assessed following the GRADE risk assessment factors(41). The lack of pooled effect size of modelling studies did not warrant us to perform a methodological diversity(42). Regarding the second objective however, variability was assessed by directly observing confidence intervals on plotted graphs.

### Data Analysis

Review findings were synthesised thematically. The quality of studies was critically appraised using most recent tools based on study design, in accordance with PHO MetQAT 1.0 quality appraisal tool(43, 44). The methodology and risk of bias of modelling studies was assessed using Relevance and Credibility Assessment (RCA) tool proposed by Caro and colleagues(45). Cohort studies were assed using Critical Appraisal Skills Programme (CASP) tool(46). Specialist Unit for Review Evidence (SURE) tool was used to assess cross sectional studies(47). Studies were grouped into 6 main categories according to outcome, as outlined in the methodology section for easy analysis and synthesis. Quality of evidence generated by different studies was assessed using the Grading Recommendations Assessment, Development and Evaluation (GRADE) tool(48).

### Data Presentation and Visualization

Tabular and graphical methods were deployed in presenting results. The GRADE summary of findings table was used to present certainty of evidence and a bar chart to present the effect direction of studies for the primary objective. In the secondary objective, forest plots were used to present the proportion of asymptomatic cases of SARS-CoV-2, using an excel model proposed by Neyeloff et al(91)

### Criteria for Prioritizing Results

In relation to the primary question, results of studies that evaluated the effectiveness of the intervention and control within the UK, with low risk of bias were prioritized because this was in line with the review objective. Real-time studies were also prioritized as these are more likely to be close to reality.

## Selection and Publication Bias

Preferential publication was counteracted by including grey literature. Missing data effect verification was performed by searching for grey literature that sought to compare the effectiveness of the intervention to the control(49). Editorials, thesis, protocols and news articles were excluded. This increased quality of included articles.

## Search Results

The search yielded 286 articles from Google Scholar, 20 articles from Science Direct, 14 articles from Mendeley, 27 articles from Pubmed and 15 articles from other sources giving a total of 362 articles. Altogether 64 eligible articles were screened for inclusion. Given the ambiguity in the use of contact tracing in most studies to include testing, studies evaluating the effectiveness of contact tracing provided they had a component of mass testing were included. Considering the novelty of the term Test and Trace used in this study, it is common place to find contact tracing based on symptom testing used in studies to be likened to Test and Trace in this review.

A total of 35 articles that met the eligibility criteria were included. Table 1 below shows summary characteristics of included studies. A flow chart of how articles were selected can be seen in figure 3 below. Summary characteristics of studies excluded due to eligibility criteria are presented in supplement 1 of the appendix.

**Figure 3:**
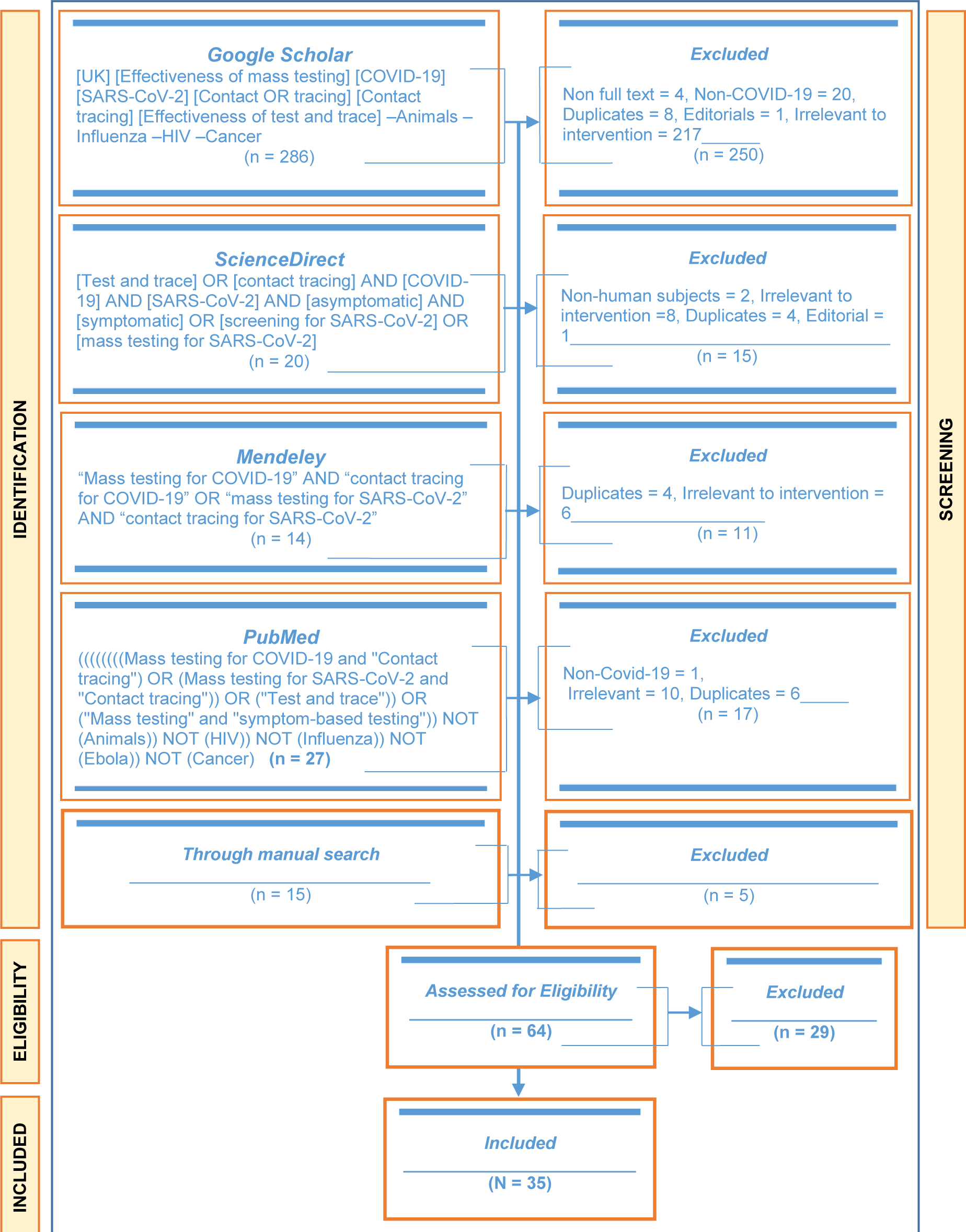
Study flow Chart: Showing n articles at each stage with N articles included in the review

**Table 1.** Summary characteristics of included studies

| Study | Design/Outcome | Sample/setting | Strategies | Assumptions | Measurements or parameters | Analysis | Findings |
| --- | --- | --- | --- | --- | --- | --- | --- |
| **1. Primary Question** |  |  |  |  |  |  |  |
| **Effectiveness (Outcome 1)** |  |  |  |  |  |  |  |
| Emery(2020) (23)(http://www.ncbi.nlm.nih.gov/pubmed/32831176) | Modelling study<br>Transmissions from asymptomatic carriers | 3,711 cruise ship passengers & crew<br>Japan | - Symptom-based testing<br>- Symptom-agnostic testing | - Constant infectiousness<br>- Progress to presymptomatic/asymptomatic is irrespective of origin of infections<br>- Unavailable symptom onset date for 115 cases proportional to cases with reported dates<br>- Unavailable test dates for 13 persons proportionate to tests among those with unreported symptom onset<br>- Proportion of asymptomatic infections and infectiousness<br>- Individual test negative after infectious period<br>- Test accuracy = 100%<br>- People are 50% more likely to be tested in biased symptom-agnostic testing | - Asymptomatic infections<br>- Presymptomatic infections<br>- Relative passenger-crew contact rate<br>- Latent period duration<br>- Duration of asymptomatic infection<br>- Age dependent proportion asymptomatic | R version 3.5.0 and LibBi Bayesian calibration with RBi | Testing irrespective of symptoms showed to be more effective in case identification than symptom-based testing |
| Grassly(2020)(50)(https://www.thelancet.com/action/showPdf?pii=S1473-3099%2820%2930630-7) | Modelling study<br>% reduction in reproduction number R | Modelled sample,<br>UK | - Symptom-based self-isolation<br>- Symptom testing and case isolation<br>- Regular testing of high-risk groups irrespective of symptoms & isolation<br>- Test and trace of contacts & isolation<br>- Contact tracing by symptoms alone | - Asymptomatic individuals are less infectious than symptomatic individuals<br>- 100% PCR test sensitivity<br>- 100% coverage of Test and Trace<br>- Sample collection done at symptom onset<br>- 1 day delay from sample collection and quarantining of contacts<br>- 80% of symptomatic cases are reported | - Proportion of asymptomatic infections<br>- Relative infectiousness<br>- Average serial interval if no isolation<br>- Incubation period<br>- PCR test sensitivity over infection diagnosis interval | Simulations | Self-isolation upon symptom onset will reduce transmissions by 47% (95% Uncertainty Interval, UI: 32-55) Screening all healthcare workers and other high at-risk populations every week will further reduce transmission by 23% (95% Uncertainty Interval: UI 16-40) in addition to that achieved by isolation (if PCR test results return within 24 |
|  |  |  | - Test-trace-test contacts and isolate | - 80% of symptomatic contacts are traced<br>- Testing done the day of symptom onset |  |  | hours), while Test and trace will further reduce transmissions by 26% (95% UI:14-35) |
| Tsou(2020)(51) (https://www.sciencedirect.com/science/article/pii/S1551714420301798) | Modelling study<br>COVID-19 outbreak containment | 393 by April 13, 2020,<br>Taiwan | A) Symptom-based testing and isolation of index cases<br>B) Mass testing of symptomatic and asymptomatic subclinical cases<br>C) Symptom-based testing, isolation and quarantine of all at risk group | - Incubation period per case and symptom onset to isolation delay follows a Weibull distribution<br>- Potential secondary cases follow a negative binomial distribution with mean = reproduction number R<br>- Strategies differed in their control of subclinical cases<br>- Initial number of cases = 5, 20 & 40<br>- At-risk persons investigated = 40%, 60%, 80% & 90%<br>- 40%, 60%, 80% of subclinical cases assumed to be detected and isolated<br>- Subclinical cases can completely be prevented | - % of subclinical cases<br>- Epidemiological investigations<br>- isolation effectiveness<br>- incubation period<br>- Number of initial cases<br>- Number of secondary infections per new infection<br>- Serial interval | Simulations | Symptom-based testing, isolation and quarantining all subclinical cases was most effective<br>However, strategy B was better than A in the prevention of transmissions prior to symptom onset |
| Mizumoto(2020)(52) (https://www.eurosurveillance.org/content/10.2807/1560-7917.ES.2020.25.10.2000180) | Modelling study<br>Asymptomatic proportion | 3,063 cruise ship passengers<br>Japan | Mass testing | N/A | - Number of test<br>- Positive cases<br>- Asymptomatic cases<br>- Symptomatic cases | Hamiltonian Monte Carlo (HMC) with the No-U-Turn-Sampler (NUTS) | A total of 634 cases were detected 328 of whom were asymptomatic.<br>Proportion of asymptomatic increased over the weeks |
| Sasmitha(2020)(53) (https://ghrp.biomedcentral.com/articles/10.1186/s41256-020-00163-2) | Modelling study<br>- Incidence<br>- Peak of cumulative COVID-19 | COVID-19 daily data<br>Indonesia | Scenario 1 = $u_1+u_4+u_5$<br>Scenario 2 = $u_1+u_2+u_4+u_5$<br>Scenario 3 = $u_1+u_2+u_3+u_4+u_5$ | - 95% false positive rate from susceptible to exposed<br>- Reinfection possibility as recovered patients loss immunity<br>- All parameters assumed to be positive and constant | - Number of susceptible persons<br>- Number of exposed individuals<br>- Number of SARS-CoV-2 carriers<br>- Number of reported infectious cases | MATLAB and R software | COVID-19 cases attained peak for strategy 1, 2 and 3 on 59<sup>th</sup>, 38<sup>th</sup> and 40<sup>th</sup> day after initial outbreak with 33151, 37908 and 39305 cases respectively |
|  |  |  | U1 = Large-scale social restriction<br>U2 = Contact tracing<br>U3 = Mass testing<br>U4 = Case detection and treatment<br>U5 = Face masks use | - Availability of rapid PCR tests | - Number of recover cases<br>- Infection rate between the susceptible and infectious persons<br>- Immunity rate |  | The optimal control measure is scenario 2 with (u1), (u2), (u4) and (u5) (see column 4) |
| Moghadas(2020)(54)(http://www.ncbi.nlm.nih.gov/pmc/articles/PMC7395516/) | Modelling study<br>Required isolation Curtail of silent transmission | 10,000 hypothetical population<br>Canada | - No self-isolation<br>- 100% severe cases self-isolate<br>- 100% symptomatic case self-isolate<br>- 100% isolation of symptomatic cases plus detection and isolation of asymptomatic cases | - Proportion of asymptomatic infections is 17.9% and 30.8% | - Secondary cases at each stage<br>- Proportion of the attack rate attributable to asymptomatic infection<br>- Proportion of the attack rate attributable to presymptomatic infection<br>- Proportion of the attack rate attributable to symptomatic infection | Simulations<br>Agent Based Model Lab | Isolating all symptomatic will still be inefficient in outbreak control<br>Combined with case isolation, our results indicate that 33% and 42% detection and isolation of silent infections would be needed to suppress the attack rate below 1%, for asymptomatic proportions of 17.9% and 30.8%, respectively |
| Bracis(2020)(55)(https://linkinghub.elsevier.com/retrieve/pii/S2468042720300737) | Modelling study<br>SARS-CoV-2 transmission projection | Daily cases of King County from Mar 8-Mar 29<br>USA | - No intervention<br>- Isolating the elderly<br>- Schools opening in fall<br>- Testing, treatment, isolation and contact tracing in combination with physical distancing | - 20% of infections are symptomatic<br>- Homogenous infectivity and outcome<br>- Constant diagnostic rate during reopening<br>- > 40% diagnosed during early testing<br>- 50% of contacts are successfully traced<br>- Contact tracing permits 5% of asymptomatic and subclinical to be tested<br>- Differential post COVID-19 physical interaction | - Non-infectious exposed<br>- Status of infection<br>- Asymptomatic cases<br>- Subclinical cases<br>- Symptomatic cases<br>- Recovered<br>- Treatment status<br>- Hospitalized | NSGA-II multivariate optimization algorithm in the mco R package<br>Monté Carlo | Ramping up testing, isolation and contact tracing of symptomatic cases reduced post-COVID interactions by 60% and very few deaths<br>Mass testing was not found to be feasible |
| Pollmann(2020)(56)(https://doi.org/10.1101/2020.05.14.20101010) | Modelling study | 10,000 hypothetical | - Digital contact tracing (based on | - All contacts using digital contact tracing can be traced | - Fraction of exposed population | Monté Carlo with second order | Contact tracing must be combined with either random mass testing or |
| 1101/2020.09.13.20192682) | Impact of digital contact tracing | al population | reported symptoms),<br>- Quarantining,<br>- Testing<br>- Social distancing<br>- Random testing | - Unreported symptoms and untested symptomatic cases<br>- Tracing of infected contacts<br>- Immediate quarantine upon report of symptoms<br>- 100% test accuracy<br>- Homogeneous population<br>- Immunity once recovered<br>- No symptoms-testing delay<br>- Absence of manual tracing<br>- Fixed latent period<br>- Backward/Forward tracing | - Fraction of sick population<br>- Population in quarantine<br>- Effective reproduction number<br>- Fraction of simulations with no outbreak |  | social distancing to control epidemic<br>Daily random testing of 20% of population as effective as social distancing |
| Hill(2020)(57)(https://doi.org/10.1101/2020.10.15.20208454) | Modelling study<br>Reduction in infections | 1,000 hypothetical population, using 2010 Social contact survey data<br>UK | - Test and trace<br>- Regular mass testing | - Each person can infect<br>- Contact network follows poisson distribution<br>- Contact probabilities fall with level of accommodation<br>- No random accommodation<br>- People can infect 1 day post symptom onset<br>- 100% test specificity<br>- Possible to Forget contacts<br>- Self-isolation time=10 days<br>- Test- results delay= 2 days<br>- Contact isolation = 14 days<br>- Adherence to test and trace<br>- No contacts during isolation<br>- No COVID-19 student beginning the term | - Timing and frequency of mass testing<br>- Coverage<br>- Adherence to isolation, test and trace<br>Proportion infected<br>- Time spent in isolation | Simulations | Daily and weekly testing combined with contact tracing adherence reduced the number of infections by more than 50% compared to test and trace alone |
| Gorji(2020)(58)(https://doi.org/10.1101/2020.03.27.20045237) | Modelling study<br>Reduction in reproduction number | Switzerland | - Mass testing<br>- Contact tracing<br>- Smart testing<sup>2</sup> and contact tracing | - 90% infection reduction due to self-isolation<br>- Basic reproduction number of 2.4 if no mitigation<br>- Test results take 1 days<br>- Children under 10 contribute little to infections<br>- At-risk subpopulation have 27 fold prevalence rate | - Number of required test<br>- Reproduction number<br>- Population using app<br>- Identified individuals with high contacts<br>- Tested population<br>- Number of contacts<br>- Contact traced | MATLAB and the Statistics Toolbox Release 2018b | Testing high risk individuals irrespective of symptoms with contact tracing will reduce R to 1.<br>Contact tracing based on symptom testing will miss most cases. |

<sup>2</sup> Mass testing of individuals with high contact rates (at-risk group)

| Study | Design/Outcome | Sample/setting | Strategies | Assumptions | Measurements or parameters | Analysis | Findings |
| --- | --- | --- | --- | --- | --- | --- | --- |
|  |  |  |  | - Detection of high contact individuals every 7 days |  |  |  |
| Alsing(2020) (59)(https://doi.org/10.1101/2020.05.05.20092221) | Modelling study<br>Efficacy | 2011 commuter data BBC pandemic dataset<br>1000 simulations<br>UK | - Contact tracing and social distancing<br>- Contact tracing with Mass testing<br>- Contact tracing with lockdowns | - Active infections at 8 months<br>- Number of daily tests required<br>- Effective reproduction number ($R_E$) per scenario<br>- number of people in lockdown | - Social interaction at home, school, work and community<br>- Offsprings drawn from a negative binomial distribution with mean=$R$<br>- Reproduction number ($R$) is country specific<br>- Herd immunity<br>- Baseline $R_0 = 2.5$<br>- Onset-to-isolation delay follows a Weibull distribution with mean 3.43 and 2.4 days standard deviation<br>- Generation time is normally skewed | - | Possible to control 38% of outbreak simulations within 8 months using contact tracing with 63.3% of outbreak still leaves $R > 1$<br>Mass testing and contact tracing contains 74% of the outbreak simulations with 36.8% of outbreaks results in $R < 1$ |
| Hagan(2020) (60)(https://www.cdc.gov/mmwr/volumes/69/wr/mm6933a3.htm) | Cross sectional<br>Prevalence of SARS-CoV-2 | 16,161 incarcerated persons in six jurisdictions<br>USA | - Symptom-based testing<br>- Mass testing |  | - Number of positive cases per test approach | - | Mass testing increased the number of COVID-19 cases from 642 (range = 2–181, median = 19) after symptom-based testing to 8,239 (range = 10–2,193, median = 403) giving a median increase of 12.3 fold |
| **Cost-Effectiveness (Outcome 2)** |  |  |  |  |  |  |  |
| Paltiel(2020) (61)(https://jamanetwork.com/journals/jamanetworkopen/fullarticle/2768923?utm_campaign=article) | Modelling study<br>Clinical Performance (tests, required isolation and infections)<br>Economic performance | 4,990 hypothetical cohort<br>USA | - Base case scenario with a reproduction number ($R_t$) of 2.5, test specificity of 98% and 10 new infections each week<br>- Worst case scenario with an $R_t$ of 3.5, test specificity of 98%, | - Test frequency = 1, 2, 3 & 7<br>- Test sensitivity = 70%- 99%<br>- Importation of infections via exogenous shocks<br>- Specificity of 98% - 99%<br>- Reproduction number = 1.5, 2.5 and 3.5<br>- Case fatality = 0.05%<br>- 30% chance that infection will lead to virus symptoms<br>- Cost per test = \$10 - \$50<br>- Abbreviated 80-days period | - Number of test administered<br>- Number of true-positives<br>- Number of false positives<br>- Number of new infections<br>- Required person-days for isolation<br>- Cost<br>- Incremental cost-benefit | Microsoft Excel with an interactive dashboard | A willingness-to-pay of $\leq \$5,500$/infection averted, screening every week using a 70% sensitive test was optimal. Regular screening (7, 3 & 2 days) was optimal if only a single test of \$25 with 80% sensitivity was available<br>There was no condition under which symptom-based |
| | (cost, incremental cost and budget) | | and 25 new infections every week<br>- Best case scenario with an Rt of 1.5, test specificity of 99.7%, and 5 new infections each week | - A cohort of non-immune students in congregate setting of 5,000 students<br>- 8-hour test turnaround time<br>- Availability of 100% confirmatory tests at \$100 | - Budget impact | | screening alone will contain outbreak |
| **Safety (Outcome 3)** |  |  |  |  |  |  |  |
| None |  |  |  |  |  |  |  |
| **Acceptability (Outcome 4)** |  |  |  |  |  |  |  |
| None |  |  |  |  |  |  |  |
| **Equity (Outcome 5)** |  |  |  |  |  |  |  |
| None |  |  |  |  |  |  |  |
| **2. Secondary Question** |  |  |  |  |  |  |  |
| **Proportion of Asymptomatic Cases (Outcome 6)** |  |  |  |  |  |  |  |
| Nishiura(2020)(62)([https://www.ijidonline.com/article/S1201-9712(20)30139-9/pdf](https://www.ijidonline.com/article/S1201-9712%2820%2930139-9/pdf)) | Cross sectional<br>Asymptomatic ratio | 565 passengers<br>Japan | RT-PCR testing | N/A | - Positive cases<br>- Asymptomatic cases<br>- Symptomatic cases |  | 63 passengers were symptomatic.<br>Four (30.8%, 95% CI: 7.7–53.8%) of 13 positive cases were asymptomatic and 9 were symptomatic |
| Porru(2020)(63)(https://www.mdpi.com/1660-4601/17/14/5104) | Cohort study<br>Health surveillance | 5,942 staff of a large hospital<br>Italy | Mass RT-PCR testing using oropharyngeal and nasopharyngeal swabs | N/A | - Positive cases<br>- Asymptomatic cases<br>- Symptomatic cases | Fisher's exact test or the chi-square test ANOVA or the Kruskal–Wallis test | A total of 238 cases detected of whom 109 were asymptomatic<br>Mass testing permitted prompt isolation and monitoring of cases |
| Treibel(2020)(64)(https://www.ncbi.nlm.nih.gov/pmc/articles/PMC7206444/) | Cross sectional<br>Asymptomatic carriers | 400 healthcare staff<br>UK | Nasopharyngeal swap PCR test | N/A | - Positive cases<br>- Negative cases<br>- Symptomatic cases<br>- Asymptomatic cases |  | Twelve (27%) of 44 positive cases were asymptomatic<br>Positive<br>Fifty staff self-isolated as a result of symptoms |
| Abeysuriya(2020)(65)(https://doi.org/10.1016/j.ejogrb.2020.07.035) | Diagnostic test<br>Prevalence of SARS-CoV-2 | 180 pregnant women (39 weeks)<br>UK | Nasopharyngeal swap PCR test | N/A | - Positive cases<br>- Number asymptomatic | Medcalc Diagnostic Test Evaluation Calculator | Seven women tested positive with 6 (85.7 %, 95% CI: 42.1–99.6) as asymptomatic.<br>Symptom-based testing sensitivity was 14.3% (0.36–57.87) and specificity was 91.86% (86.72–95.48) |
| Brown(2020)(66)(https://www.sciencedirect.com/science/article/pii/S0163445320304503) | Cross section study<br>SARS-CoV-2 prevalence | 1152 healthcare workers in 6 hospitals<br>UK | Nasopharyngeal/oropharyngeal swap PCR tests | N/A | - Positive test<br>- Asymptomatic carriers<br>- Symptomatic cases<br>- Viral load | Stata (version 15, StataCorp, Texas) | Thirteen (57%) of 23 positive cases had symptoms compliant with COVID-19, of whom 4 (17.4%) were asymptomatic |
| Graham(2020)(67)(https://www.sciencedirect.com/science/article/pii/S0163445320303480) | Cross sectional<br>Infections Clinical features Outcome | 464 residents & staff in Care homes<br>UK | - Comprehensive testing with Oropharyngeal and nasopharyngeal swaps<br>- Symptom screening | N/A | - Positive case<br>- Negative cases<br>- Symptomatic cases<br>- Asymptomatic cases | Chi Square and non-parametric test in R version 3.6.0 | 126 (40%, 95% CI 35 to 46) of the 313 tested residents were positive.<br>Only 72 (57%, 95% CI 49–66) positive cases would have been diagnosed based on symptom-testing |
| Arons(2020)(68)(http://www.nejm.org/doi/10.1056/NEJMoa2008457) | Cross section survey<br>Transmission<br>Adequacy of symptom-based screening | 89 residents of skilled nursing home<br>USA | Point prevalence testing with RT-PCR on nasopharyngeal and oropharyngeal swabs | N/A | - Positive cases<br>- Negative cases<br>- Number asymptomatic<br>- Number symptomatic | SAS software, version 9.4 (SAS Institute) | 48 (63%) of 76 tested residents were positive of whom 27 (56%) were asymptomatic. 24 of the 27 developed symptoms 1 week post test |
| Jameson(2020)(69)(https://doi.org/10.1017/ice.2020.361) | Cross sectional<br>Asymptomatic infections | 121 non-symptomatic healthcare staff<br>USA | - Universal testing<br>- Universal symptom: screening<br>- Isolation of cases (RT-PCR test on nasopharyngeal swaps) | N/A | - Positive cases<br>- Negative cases<br>- Number asymptomatic<br>- Number symptomatic | - | No positive case was found among 121 out of 499 eligible healthcare workers screened |
| Callaghan(2020)(70)(https://www.cdc.gov/mmwr/volumes/69/wr/mm6926a4.htm?s_cid=mm6926a4_w) | Cross sectional<br>Effectiveness of preventive procedures & prevalence of SARS-CoV-2 | 46 patients & 171 healthcare professionals<br>US | Nasopharyngeal swap PCR test | N/A | - Positive cases<br>- Number asymptomatic | - | No participant tested positive for COVID-19 |
| Louie(2020)(71)(https://academic.oup.com/cid/advance-article/doi/10.1093/cid/ciaa1020/5873783) | Cross-sectional survey<br>Transmission monitoring | 734 persons<br>USA | Outbreak response mass testing with PCR on nasopharyngeal swaps | N/A | - Number symptomatic cases<br>- Number of asymptomatic cases<br>- Number of hospitalized cases<br>- Number of dead cases | ABI 7500 platform.<br>Automated Abbott m2000 RT-PCR platform | Mass testing identified a high proportion of asymptomatic cases. Symptom-based screening is ineffective in detecting cases among healthcare workers |
| Gudbjartsson(2020)(72)(https://www.ncbi.nlm.nih.gov/pmc/articles/PMC7175425/) | Cross sectional<br>SARS-CoV-2 transmissions | 22,279<sup>3</sup> At-risk group and general population<br>Iceland | - Targeted testing<br>- Open invitation screening<br>- Random invitation screening<br>Using nasopharyngeal and oropharyngeal samples | N/A | - Positive cases<br>- Negative cases<br>- Symptomatic cases<br>- Asymptomatic cases | R Package (https://cran.r-project.org/package=binom) | 13.3% tested positive in targeted testing, 0.8% in open invitation testing and 0.6% in random invitation testing. |
| Reid(2020)(73)([file:///E:/te lechargemen](https://www.echocare.com)) | Cross sectional | 5,204 healthcare staff | - Symptomatic testing | N/A | - Positive cases per regime<br>- Symptomatic cases | SAS statistical | 188 (6.4%) positive cases detected during symptomatic testing and 5 (0.2%) |

<sup>3</sup> Sample of 9199 targeted persons, 10797 openly invited and 2283 randomly invited

| Study | Design/Outcome | Sample/setting | Strategies | Assumptions | Measurements or parameters | Analysis | Findings |
| --- | --- | --- | --- | --- | --- | --- | --- |
| t/jammi-2020-0027.pdf) | Test uptake<br>Test positivity | Canada | - Asymptomatic testing<br>Using nasopharyngeal swabs |  | - Asymptomatic cases | software (version 9.4) | positive cases during asymptomatic testing, with a low probability of testing positive |
| Lavezzo(2020)(74)(https://www.nature.com/articles/s41586-020-2488-1?fbclid=IwAR0Y69FXQqJqogOf-1ln) | Cross sectional<br>Asymptomatic infection contribution | 2,812<br>2,343<br>Italy | RT-PCR on nasopharyngeal swabs | N/A | - Number of positive cases per survey<br>- Number of negative tests per survey<br>- Number of positive cases that are asymptomatic per survey | Fisher's exact test<br>Exact<br>Wilcoxon–Mann–Whitney test | The first survey gave a prevalence of 2.6% (95% CI: 2.1–3.3%) and 1.2%; 95% CI: 0.8–1.8%) for survey 2. 29 (39.7%; 95% CI: 28.5–51.9%) of positive tests in the survey 1 were asymptomatic and 13 (44.8%; 95% CI: 26.5–64.3%) in survey 2. |
| Kimball(2020)(75)(http://www.cdc.gov/mmwr/volumes/69/wr/mm6913e1.htm?s_cid=mm69) | Cross sectional<br>Utility of symptom screening | 76 older adults in skilled nursing care home<br>USA | RT-PCR mass testing on nasopharyngeal and oropharyngeal swabs | N/A | - Positive cases detected<br>- Asymptomatic cases<br>- Symptomatic cases | SAS statistical software (version 9.4) | Twenty three (30%) residents were positive with 13 (57%) either presymptomatic or asymptomatic. Testing based on symptom screening could miss up to 50% of cases |
| Olalla(2020)(76)(https://academic.oup.com/qjmed/article/113/11/794/5890491) | Cross sectional<br>Prevalence of asymptomatic cases | 498 healthcare workers<br>Spain | - Symptom screening<br>- Asymptomatic testing<br>PCR on naso- and oropharyngeal swabs | N/A | - Positive cases<br>- Positive IgG and IgM<br>- Number symptomatic on day of sampling<br>- Number symptomatic 14 days prior to sampling | SPSS 15, | 2 asymptomatic on day of sampling tested positive. 1 reported having gad symptoms in last 14 days |
| Guery(2020)(77)(https://www.sciencedirect.com/science/article/pii/S0399077X20301268) | Cross sectional<br>SARS-CoV-2 cases | 136 nursing care home staff<br>France | RT-PCR mass testing (using Nasopharyngeal swap) | N/A | - Positive cases detected<br>- Asymptomatic cases<br>- Symptomatic cases | - | Three (2.2%) cases detected, 1 of which was symptomatic and 1 developed symptoms within 24 hours |
| Roxby(2020) (78)(http://www.cdc.gov/mmwr/volumes/69/wr/mm6914e2.htm?s_cid=mm) | Cross sectional<br>COVID-19 morbidity | 142 staff & residents in residential community<br>USA | RT-PCR mass testing (using Nasopharyngeal swap) | N/A | - Positive cases detected<br>- Asymptomatic cases<br>- Symptomatic cases | - | Five (7%) cases were detected, 3 were asymptomatic<br>Symptom-based testing might not identify all positive cases |
| Lytras(2020) (79)(a)(https://academic.oup.com/jtm/article/doi/10.1093/jtm/taaa054/5820895) | Cross sectional<br>SARS-CoV-2 prevalence | 357 passengers repatriated from UK<br>Greece | RT-PCR mass testing (using Nasopharyngeal swap) | N/A | - Positive cases detected<br>- Asymptomatic cases<br>- Symptomatic cases | - | Thirteen (3.6%, CI: 2.0–6.1) positive asymptomatic cases |
| Lytras(2020) (79)(b)(https://academic.oup.com/jtm/article/doi/10.1093/jtm/taaa054/5820895) | Cross sectional<br>SARS-CoV-2 prevalence | 394 passengers repatriated from Spain<br>Greece | RT-PCR mass testing on nasopharyngeal swaps | N/A | - Positive cases detected<br>- Asymptomatic cases<br>- Symptomatic cases | - | Twenty five (6.3%, 95% CI: 4.1–9.2%) positive asymptomatic cases |
| Lytras(2020) (79)(c)(https://academic.oup.com/jtm/article/doi/10.1093/jtm/taaa054/5820895) | Cross sectional<br>SARS-CoV-2 prevalence | 32 passengers repatriated Turkey<br>Greece | RT-PCR mass testing (Nasopharyngeal swap) | N/A | - Positive cases detected<br>- Asymptomatic cases<br>- Symptomatic cases | - | Two (6.3%, 95% CI: 0.8–20.8%) positive asymptomatic cases |
| Hoehl(2020)(80)(https://www.nejm.org/doi/10.1056/NEJMc2001899) | Cross sectional<br>SARS-CoV-2 infection | 125 passengers evacuated from Wuhan<br>Germany | RT-PCR mass testing (Nasopharyngeal swap and sputum) | N/A | - Positive cases detected<br>- Asymptomatic cases<br>- Symptomatic cases | - | Two (1.8%) of 114 asymptomatic passengers tested positive. All 11 symptomatic patients tested negative<br>Symptom-based testing failed to detect SARS-CoV-2 patients. |
| Cao(2020)(81)(https://www.nature.com/articles/s41467-020-19802-w?fbclid=IwAR2mBbllfSaqfobclD5cOcjQ3yh9KjTp_nNCISj-MuT3Y8GyLv-AfrB3_efY) | Cross sectional<br>Prevalence estimate | 9,899,828 residents in Wuhan<br>China | Citywide mass testing using TR-PCR on nasopharyngeal and throat swabs | N/A | - Positive cases detected<br>- Asymptomatic cases<br>- Symptomatic cases | R peackage “binom” version 1.1-1<br>SPSS version 22.0 | No symptomatic case was found compared to 300 asymptomatic cases (0.303/10,000, 95% CI 0.270–0.339/10,000) |
| Baggett(2020)(82)(https://www.ncbi.nlm.nih.gov/pmc/articles/PMC7186911/) | Cross sectional<br>Positive SARS-CoV-2 cases | 408 homeless shelter residents<br>USA | - Mass testing<br>- Symptom screening<br>(PCR on nasopharyngeal swabs) | N/A | - Positive cases<br>- Symptomatic cases<br>- Asymptomatic cases | - | 147 (36.0%) subjects tested positive, of whom 87.8% were asymptomatic |
| Imbert(2020)(83)(https://www.ncbi.nlm.nih.gov/pmc/articles/PMC7454344/pdf/ciaa1071.pdf) | Cross sectional<br>Infected persons | 150 homeless shelter residents<br>USA | Mass RT-PCR testing on nasopharyngeal specimens | N/A | - Positive cases detected<br>- Asymptomatic cases<br>- Symptomatic cases | SAS System (SAS Institute Inc., Cary, NC) | Fity two (52%) of tested residents were asymptomatic. This occurred when registered incidence was 5.1 case per 100,000 |

## Methodological and Risk of Bias Assessment

I went for whole study assessment and deployed study design specific tools due to the lack of a standardized tool for non-randomised controlled studies(44, 84). The critical appraisal is also in accordance with PHO MetQAT 1.0 quality appraisal tool(43).

### Modelling studies

A total of 12 modelling studies were included and assessed for bias, 5 (41.7%) of which were rated at low risk, 4 (33.3%) at moderate risk and 3 (25%) at high risk of bias. The main concerns for ratings are summarized in table 2 below;Cohort Studies

**Table 2.** Risk of bias of modelling studies

| Study | Relevance | Credibility | Overall Risk Assessment |
| --- | --- | --- | --- |
| **Primary question** |  |  |  |
| **Effectiveness** |  |  |  |
| Emery et al(23) | Insufficient | Insufficient | Low: Unsuitable population and setting |
| Grassly(2020)(50) | Sufficient | Sufficient | Low |
| Tsou(2020)(51) | Insufficient | Insufficient | High: Reduced at-risk population to subclinical cases. Population, setting and dataset were not appropriate. Contact tracing not considered. Validation process was not reported. Conflict of interest not declared |
| Mizumoto(2020)(52) | Insufficient | Insufficient | Moderate: Unsuitable population and setting, inadequate internal validity and treatment of structural and parameter uncertainty |
| Sasmita(2020)(53) | Insufficient | Insufficient | Moderate: Validation process unclear, Measures highlighted have been implemented in many countries (including the UK) that still see their daily cases rising. Setting not relevant |
| Moghadas(2020)(54) | Sufficient | Insufficient | High: Population and setting not relevant with some missing outcome measures. Validation process unreported. No real-life dataset was used in modelling |
| Bracis(2020)(55) | Insufficient | Sufficient | Low: Population and setting were not relevant |
| Pollmann(2020)(56) | Insufficient | Insufficient | High: Hypothetical population and setting, no use of real-world data in parameterization, possible conflict of interest, |
| Hill(2020)(57) | Sufficient | Sufficient | Low: No concerns |
| Gorji(2020)(58) | Insufficient | Insufficient | Moderate: Unrepresentative population and setting no use of real word data and declaration of conflict of interest |
| Alsing(2020)(59) | Sufficient | Insufficient | Low: Underreported validation process and undeclared conflict of interest |
| **Cost-effectiveness** |  |  |  |
| Paltiel(2020)(61) | Insufficient | Insufficient | Moderate: Unsuitable population and setting. No use of real-world dataset and neither was the validation process reported. Lack of precision |

The lone included cohort study was rated at moderate risk of bias. A summary of the methodological assessment is presented in table 3 below;

**Table 3.** Risk of bias of Cohort Studies

| Study | Risk of bias | Main concerns |
| --- | --- | --- |
| **Secondary question** |  |  |
| **Proportion of asymptomatic cases** |  |  |
| Porru(2020)(63) | Moderate | Unrepresentative population and setting, contact tracing limited to control, no eligibility criteria, unreported loss to follow-up |

### Cross sectional studies

Out of 22 cross-sectional studies assessed, 5 (23%) were judged to be at low risk of bias, 1 (4%) at moderate risk and 16 (73%) at high risk of bias. Table 4 below highlights the main concerns of bias.

**Table 4.** Risk of bias of cross sectional studies

| Study | Risk of bias | Main concerns |
| --- | --- | --- |
| **Primary question** |  |  |
| **Effectiveness** |  |  |
| Hagan(2020)(60) | High | Lack of study design, no contact tracing, unrepresentative population and setting, unjustified sample size, No eligibility criteria leading to possible selection issues, reasonable refusal rate, unreported statistical methods, actual sample collection and testing procedures unreported and unreported participant characteristics. |
| **Secondary Question** |  |  |
| **Proportion of asymptomatic cases** |  |  |
| Nishiura(2020)(62) | High | Unrepresentative population and setting. No contact tracing, unspecified study design, participant characteristics not provided, underreported statistical analysis and results, details of sample management and testing unreported, unreported study limitations |
| Treibel(2020)(64) | High | Unreported study design, no contact tracing, unjustified sample size, Not sure of fairness in participant selection due to lack of eligibility criteria, unreported statistical methods, actual sample collection and testing procedures unreported, no study limitations, and declaration of conflict of interest, unreported participant characteristics. |
| Brown(2020)(66) | Low | No contact tracing, sample size justification |
| Graham(2020)(67) | Low | No contact tracing, unreported eligibility criteria and therefore issue with selection |
| Abey Suriya(2020)(65) | Low | No contact tracing, sample collection, transportation and analysis procedures unreported |
| Arons(2020)(68) | High | Unrepresentative population and setting. Lack of justification of sample size, no eligibility criteria, exclusion of asymptomatic staff, unreported statistical methods and unreported conflict of interest |
| Jameson(2020)(69) | High | Unrepresentative population and setting, study design not clear, no eligibility criteria with unknown fairness in selection, unreported participant characteristics and statistical methods, no details of sample collection and management, lack of precision in results and reported with no study limitations |
| Callaghan(2020)(70) | High | No contact tracing, unrepresentative population and setting. Unjustified sample size, eligibility criteria not reported, unreported statistical analysis. |
| Louie(2020)(71) | Moderate | No justification for sample size, description of statistical methods unclear and no eligibility criteria |
| Gudbjartsson (2020)(72) | High | Unrepresentative population and setting, no contact tracing, unreported sample size justification, eligibility criteria and study limitations |
| Reid(2020)(73) | High | Unrepresentative population and setting, no contact tracing, no study limitations, sample collection and management unreported, undeclared conflict of interest, unreported confidence intervals |
| Lavezzo(2020)(74) | Low | Participant characteristics not provided |
| Kimball(2020)(75) | High | No clear statement of study design, no contact tracing, no sample size justification, unrepresentative population and setting, no eligibility criteria. Underreported statistical methods and funding and no confidence intervals in estimates |
| Olalla(2020)(76) | High | Partial analysis with no contact tracing, unrepresentative population and setting, unjustified sample size, underreported statistical methods, no mention of eligibility criteria and funding. |
| Guery(2020)(77) | High | No contact tracing unrepresentative population/setting, selection issues due to no eligibility criteria, unjustified sample size, unreported statistical methods, imprecise reported results |
| Roxby(2020)(78) | High | Unspecified study design, no contact tracing, likely participant selection issues due to lack of eligibility criteria, no sample size justification, unreported statistical methods and lack of precision in the reported results |
| Lytras(2020)(79) | High | No study design, no contact tracing, no sample size justification and eligibility criteria. Unreported statistical methods, Lack of eligibility criteria and unclear selection of participants, no mention of collection and transportation procedure of samples and no study limitations |
| Hoehl(2020)(80) | High | Unrepresentative population and setting, no study design, no contact tracing, no sample size justification and eligibility criteria. Unreported statistical methods, Lack of eligibility criteria and unclear selection of participants, unreported collection and transportation procedure of samples, lack of precision in results, possible conflict of interest and no study limitations |
| Cao(2020)(81) | Low | No concerns |
| Baggett(2020)(82) | High | No clear statement of study design, no contact tracing in the intervention, unrepresentative population and setting, no sample size justification and eligibility criteria. Underreported statistical methods |
| Imbert(2020)(83) | High | No statement of study design, unrepresentative population and setting, no sample size justification, lack of eligibility criteria made fair selection of participants unclear, lack of precision in results, underreported sample collection, transportation testing |

## Synthesis of Results

### Primary Question: Is there evidence that mass testing and contact tracing could suppress community spread of SARS-CoV-2 infections better than test and trace?

Vote counting was deployed as the method to synthesize results, in line with direction of effect that was used. Studies were prioritized based on their degree of bias in the reported evidence. The GRADE diagram for assessing the quality of evidence was used to grade the evidence presented by the different studies(85). GRADE summary of findings table of the different studies can be seen in Table 5 below.

#### Outcome 1: Effectiveness

Four of 12 studies (33.3%) under this outcome were at high risk of bias, three (25%) were at moderate risk of bias and five (41.7%) were rated as low. Nine studies [75%, 95% Binomal Exact CI: 42.8%-94.5%, p=0.15] including Emery (2020), Tsou (2020), Mizumoto (2020), Moghadas (2020), Pollmann (2020), Hill (2020), Gorji (2020), Alsing (2020) and Hagan (2020) were voted in favour of the intervention. Three studies [25%, 95% BE CI: 5.5%-57.1%, p=0.15] including Grassly (2020), Sasmita (2020) and Bracis (2020) showed an unfavourable direction of effect and were voted in favour of the control. The body of evidence presented by the 11 modelling studies for this outcome was downgraded by 3 levels to very low. Firstly, because studies were neither randomized control trials nor real-time studies leading to one level down. An additional 2 levels downgrading was due to serious study bias, inter-study variation, imprecision and indirectness. The evidence from the lone cross-sectional study (Hagan, 2020), was downgraded by 3 levels to “very low” as well. It was downgraded by one level because the study was not a randomized control trial. It was further downgraded by 2 levels due to methodological issues, imprecision and indirectness.

#### Outcome 3: Cost-Effectiveness

A single study found for this outcome (Paltiel, 2020), was voted in favour of the intervention. This study was at high risk of bias. The quality of evidence was downgraded by one level given that it is not a randomized control trial. Being a model based on assumptions coupled with study limitations, imprecision and indirectness, the evidence was further downgraded by 2 levels. Evidence was classed as very low.

#### Outcome 2: Safety

I found no study addressing this outcome. However, a body of literature exist regarding safety and security concerns from the public with contact tracing(86–88). Also, both nasopharyngeal and oropharyngeal swaps appear to be slightly invasive. The possible harms of mass testing have also been analysed by some authors(89).

#### Outcome 4: Acceptability

Again, no study was found regarding this outcome. Altmann and colleagues found a high level of acceptance for app-based contact tracing in their investigation across different countries including the UK(90).

#### Outcome 5: Equity

There was no study for this outcome. It remains however clear that the test and trace system is not equitable(27).

### Binomial Test and 95% Confidence Interval

A total of 13 studies were retained for the primary objective. Statistical synthesis for the primary objective was based on binomial probability test and binomial exact confidence intervals performed in StataCorp 14.2 (Texas, USA) with input. Ten of the studies favoured the intervention [76.9%: 95% Binomial Exact CI: 46.2% - 95.0%, P=0.09], with just three [23.1%, 95% BE CI: 5.0%-53.8%, p=0.09] voted in favour of the control. The above indicates that the intervention is a better strategy than the control in the control of SARS-CoV-2/COVID-19 transmissions. The probability that the above estimate is true if conventional Test and Trace programme was truly better than Mass Testing and Contact Tracing is just 9%. The 76.9% favourable direction of effect is a clear enough majority vote to say that mass test and trace is truly more beneficial.

Assuming that the true probability of both MTT and TT being equivocal is 0.5 under the null hypothesis (H0: MTT=TT), this study observed 10 votes well above the expected mean of 6.5 ± 1.803 votes. Four of 10 studies (40%) in favour of the intervention were at high risk of bias, 3 at moderate risk and 3 at low risk of bias. Twenty three percent of 13 retained studies were of representative sample and setting. Two of all 3 studies (Hill, 2020 and Alsing, 2020) implemented in the UK, voted in favour of the intervention were judged to be at low risk of bias. The effect direction plot of different studies is shown in figure 4 below.

**Figure 4:**
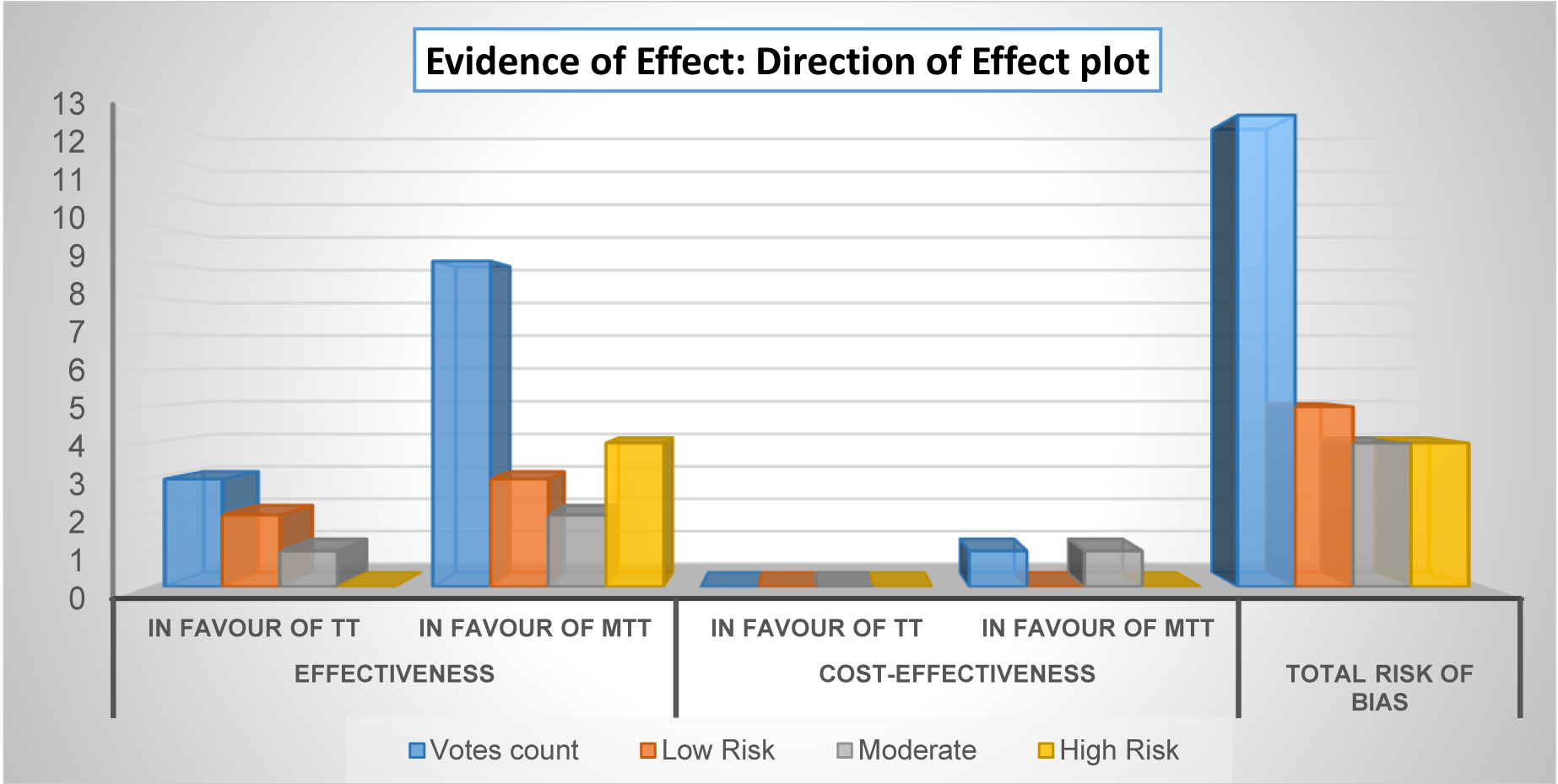
Evidence of effect attributable to the intervention and control for the primary objective. MTT = Mass Test and Trace (Intervention), TT = Test and Trace (Control)

The generated GRADE evidence profile was used to present the synthesis findings regarding the primary objective as seen in table 6 below. Supplement 6 provides details of how the evidence for different outcomes was graded.

**Table 6:**
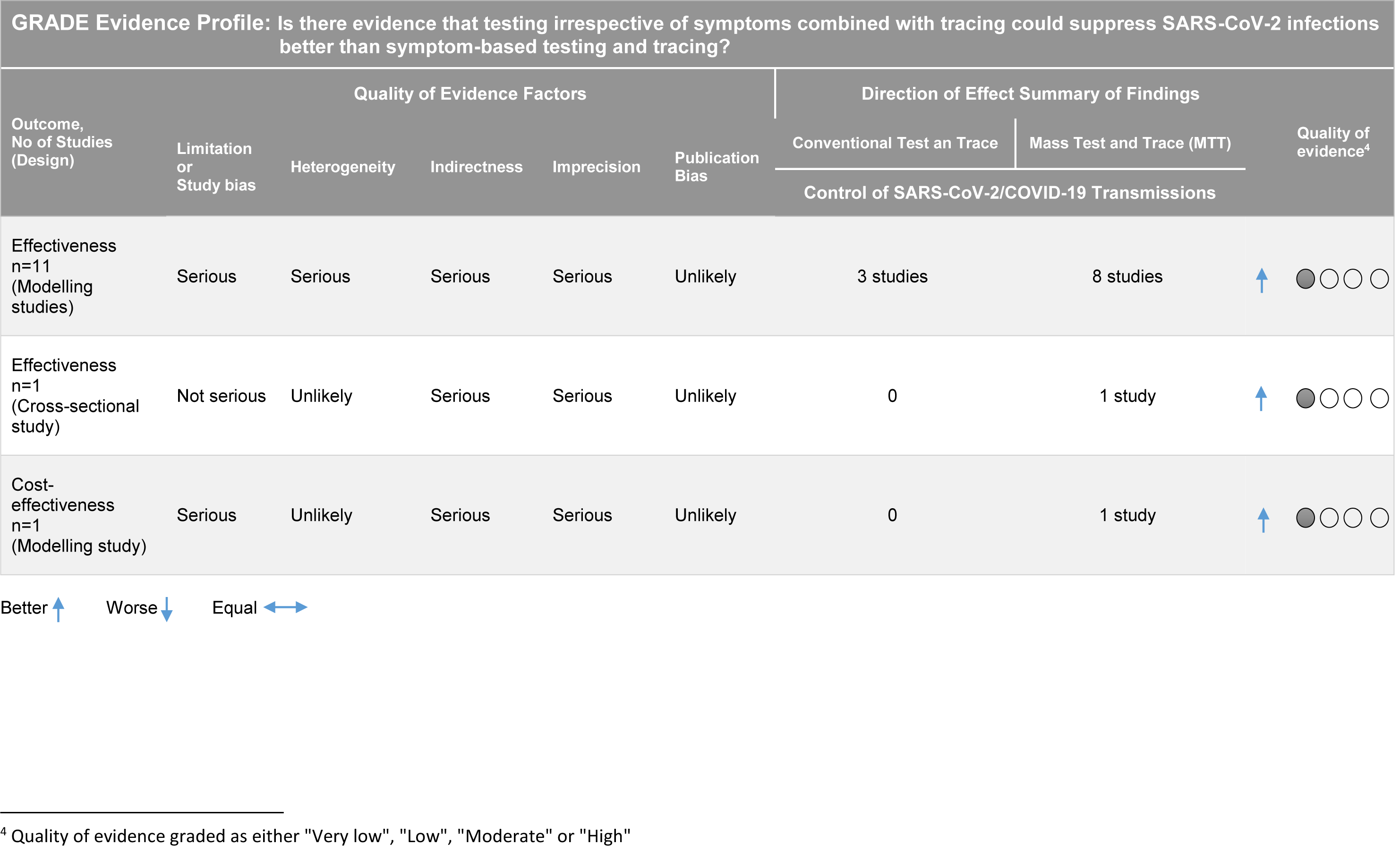
Table of Certainty of Evidence of Included Studies

The results of 6 studies including Emery (2020), Muzimoto (2020), Gorji (2020), Hill (2020), Alsing (2020) and Paltiel (2020) were prioritized. These contributed more to the conclusion that the intervention was better because they were judged to be at low to moderate risk of bias. Three of the studies (Emery, 2020; Hill, 2020; Alsing, 2020) were judged to be at low risk of bias. Two of these (Hill, 2020 and Alsing, 2020) were both of the representative population and evaluated mass testing and contact tracing as a hybrid strategy, in line with the primary objective. Emery (2020) failed to consider contact tracing but compared the effect of testing based on symptoms and testing irrespective of symptoms. The direction of effect will not be different if contact tracing were to be integrated since contact tracing is contingent on testing.

### Secondary Question: What is the proportion of asymptomatic cases of SARS-CoV-2 reported during mass testing interventions?

A total of 21 cross sectional studies and 1 cohort study (33 reports and 9942828 participants) (Nishiura, 2020; Treibel, 2020; Brown, 2020; Graham, 2020; Abeysuriya, 2020; Arons, 2020; Jameson, 2020; Callaghan, 2020; Louie, 2020; Gudbjartsson, 2020; Reid, 2020; Lavezzo, 2020; Kimball, 2020; Olalla, 2020; Guery, 2020; Roxby, 2020; Lytras, 2020; Hoehl, 2020; Cao, 2020; Baggett, 2020; Imbert, 2020) were retained under the secondary objective. Mean number of positive and asymptomatic cases were 80.6±187.7 and 32.8±57.1 respectively. There was limited precision in effect estimates with just 27% of studies providing data on confidence intervals useful for the research question. Thirty two percent of studies were at low to moderate risk of bias and 68% at high risk of bias. Risk of bias evaluation can be found in table 4 above.

### Outcome among detected positive cases

The proportion of asymptomatic cases among those testing positive ranged from 28% (95% CI: 25.9 – 30.2) in the general population to 96.6% (95% CI: 82.2 – 99.9) among care home staff. The overall proportion was found to be 40.7% (95% CI: 38.8 – 42.5) as can be seen in figure 5 below. Two studies (Jameson, 2020 and Callaghan, 2020) neither detected any cases nor found asymptomatic carriers and so were excluded.

**Figure 5:**
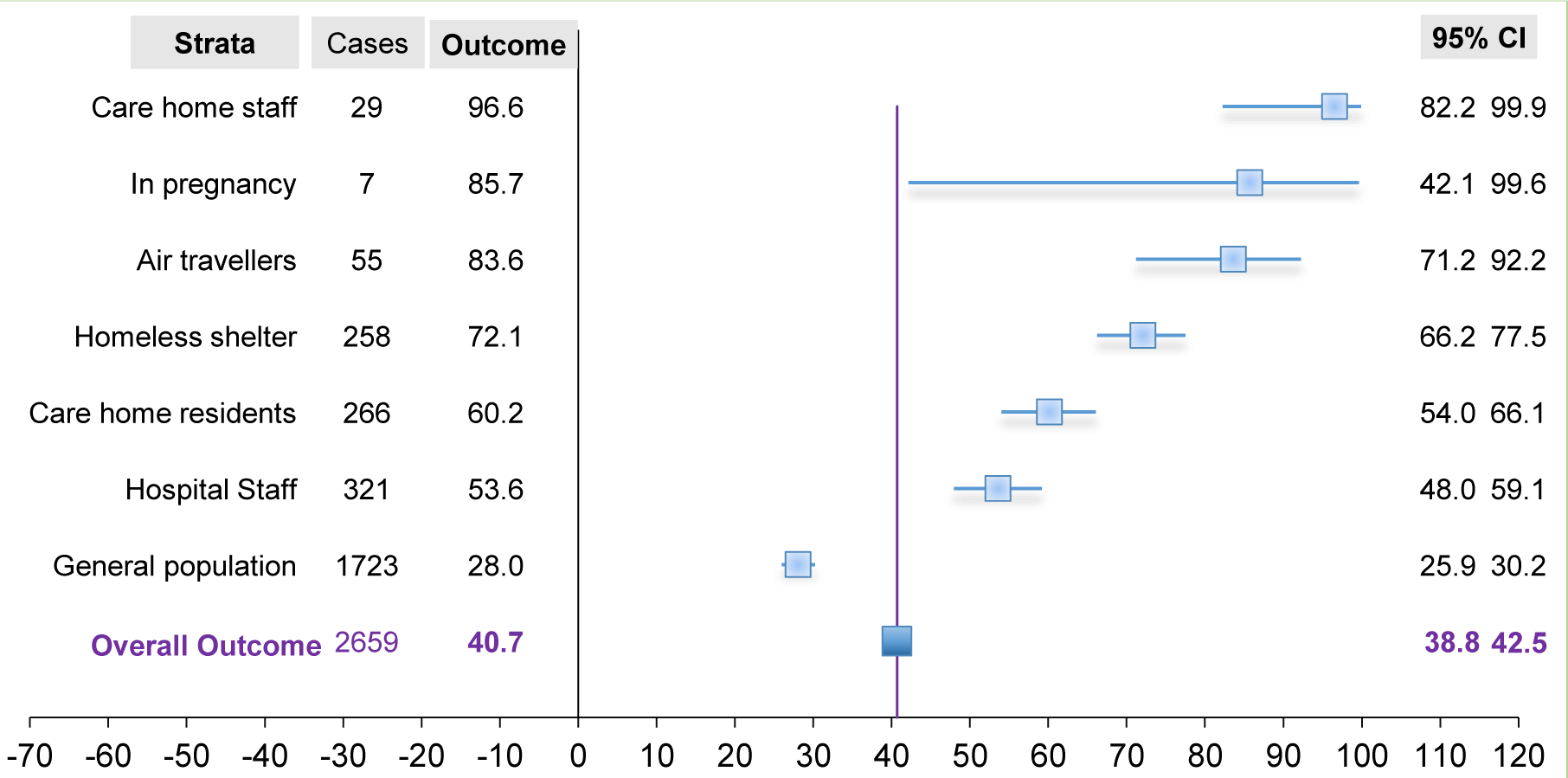
Proportion of asymptomatic SARS-CoV-2 carriers (outcome in %) among detected positive cases of SARS-CoV-2 in different settings from 22 studies CI: Confidence Interval

### Outcome in tested sample population

Prevalence of asymptomatic SARS-CoV-2 was highest among homeless shelter residents [30.1%, 95% CI: 26.5-33.9], followed by care home residents [21%, 95% CI: 18 – 24) and lowest among hospital patients [0%, 95% CI: 0.0 – 1.2]. Besides screening in the general population, overall asymptomatic SARS-CoV-2 prevalence for all other settings was 3.9% (95% CI: 3.6 – 4.2). Figure 6 below shows outcome prevalence in different specific populations.

**Figure 6:**
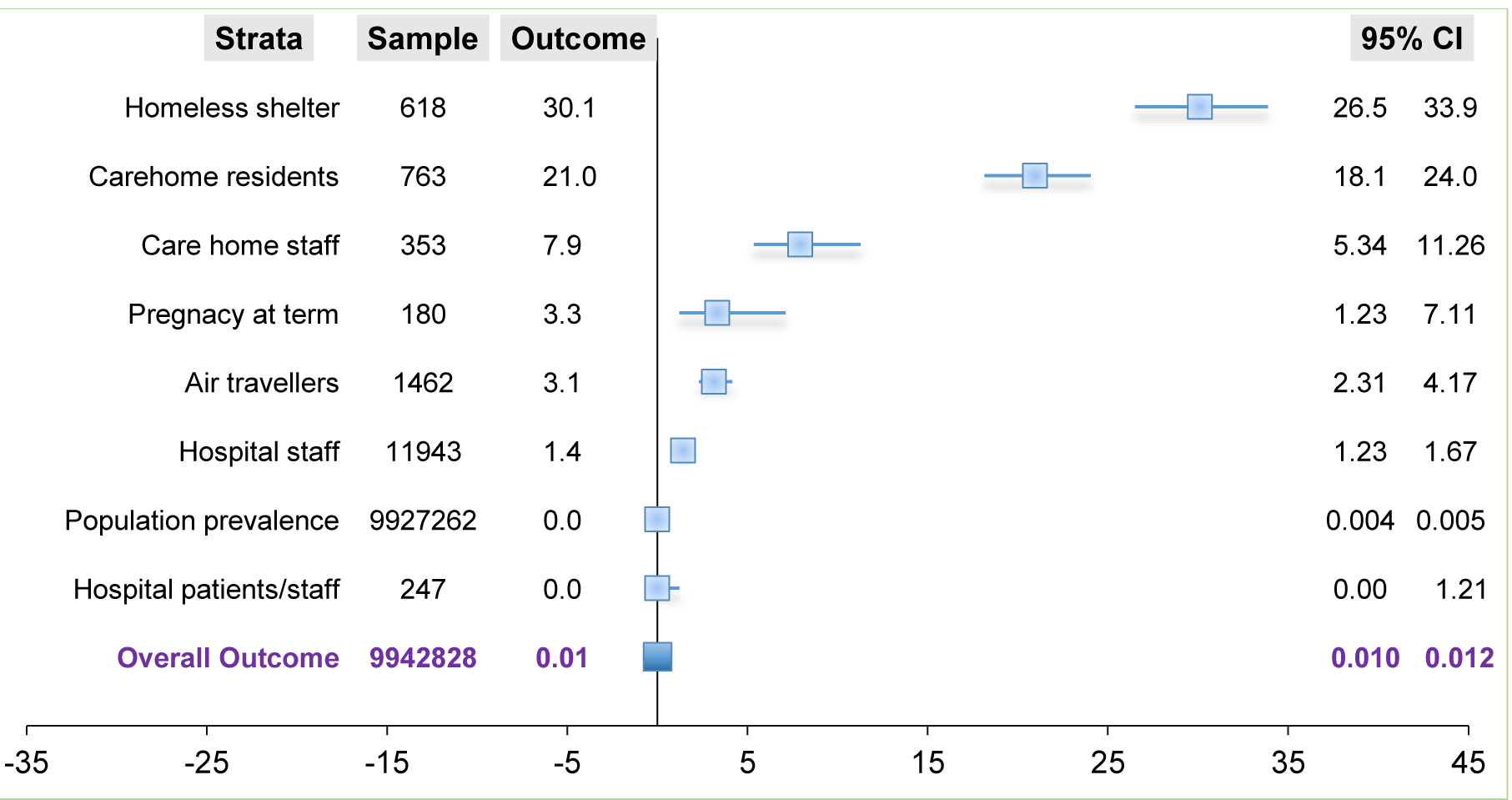
Proportion of asymptomatic SARS-CoV-2 carriers (outcome in %) in sampled population in different settings from 22 studies CI: Confidence Interval

The prevalence among asymptomatic populations from 6 studies (Louie, 2020; Treibel, 2020; Lytras, 2020; Graham, 2020; Hoehl, 2020; Reid, 2020) was 3.4% (95% CI 3-4%). The prevalence in a mixed population from 17 studies (Nishiura, 2020; Brown, 2020; Abeysuriya, 2020; Arons, 2020; Jameson, 2020; Callaghan, 2020; Gudbjartsson, 2020; Lavezzo, 2020; Kimball, 2020; Olalla, 2020; Guery, 2020; Roxby, 2020; Cao, 2020; Baggett, 2020; Imbert, 2020; Graham(a), 2020; Porru, 2020), averaged 0.01% (95% CI: 0.008-0.010). Supplement 7 and 8 provides more details.

### Outcome within the United Kingdom

Four studies including Treibel (2020), Brown (2020), Graham (2020) and Abeysuriya (2020) evaluated the outcome within the UK among hospital staff (Treibel, 2020 and

Brown, 2020), in care homes (Graham, 2020) and among pregnant women (Abeysuriya, 2020). The proportion of asymptomatic cases among those tested positive ranged from 44% (95% CI: 35.5 – 53.2) in care homes to 85.7% (95% CI: 42.1% - 100%) in pregnancy. The overall proportion among detected cases was found to be 56.6% (95% CI: 49.6 – 63.4). Figure 7 below shows the relationship of asymptomatic proportion among detected cases and in sampled population in different settings within the UK.

**Figure 7:**
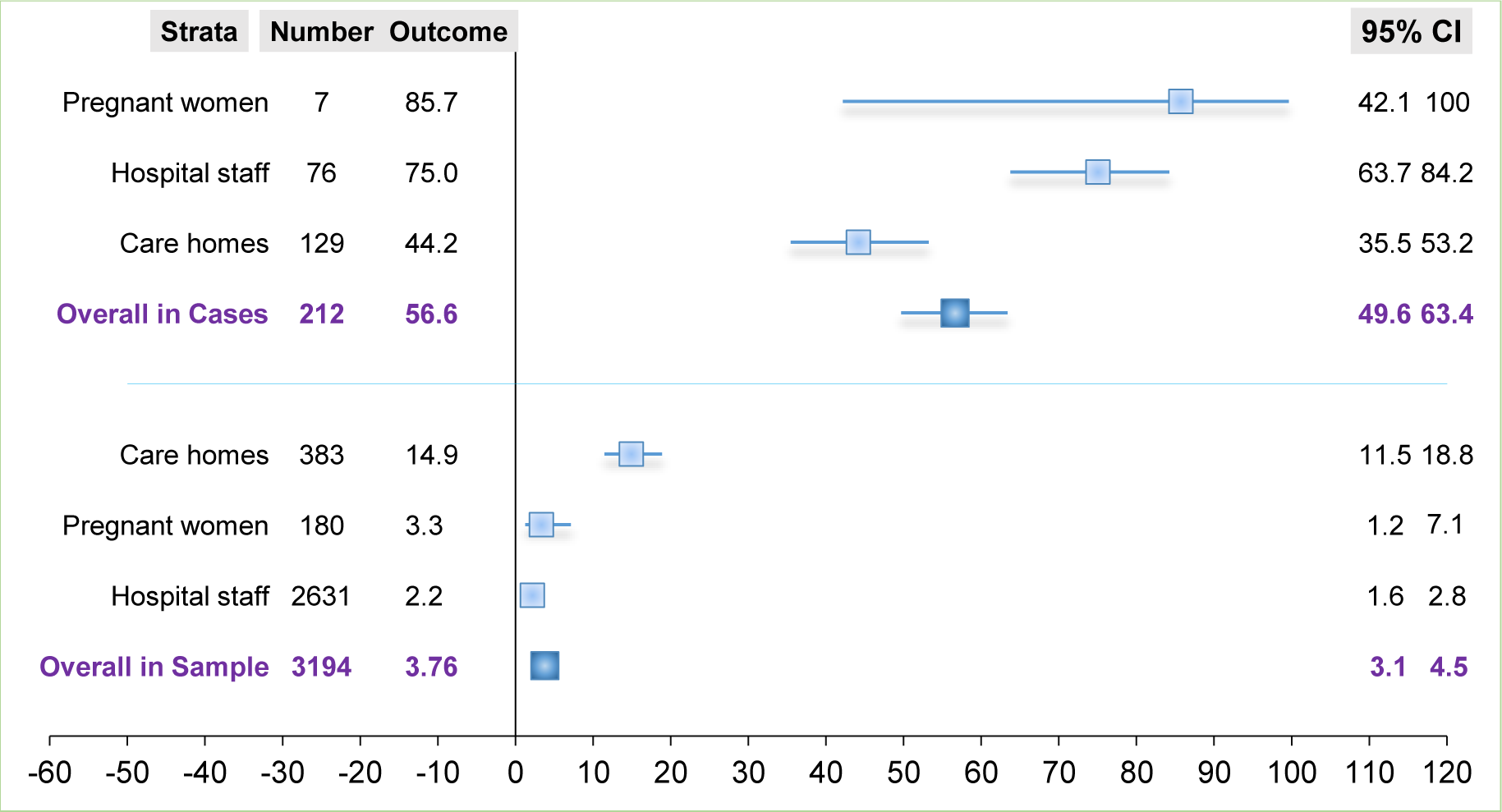
Proportion of asymptomatic SARS-CoV-2 carriers (outcome in %) in detected positive cases and sampled population in different settings from 4 studies in the UK CI: Confidence Interval

Overall prevalence of asymptomatic cases within the UK was found to be 3.76% (95% CI: 3.1 – 4.5) with rates ranging from 2.2% (95% CI: 1.6 – 2.8) among hospital staff to 14.9% (95% CI: 11.5 – 18.8) in care homes. Figure 7 above clearly demonstrates a higher overall rate among detected cases compared to that of all studies (z=4.53, p=0.00001 at p=0.05). Asymptomatic cases were 1.4 times more likely to be detected among positive cases in the UK than all studies put together. There was no significant difference between overall prevalence rate in the UK (3.76%) and all studies put together (3.9%), besides population screening (z = -0.37, p=0.71 at p=0.5)

All unreported confidence intervals were generated in SATA 14.2 (binomial exact) and exported to excel. The rule of three was applied to all studies with no outcome event (Jameson, 2020 and Callaghan, 2020).

## Inter-study Variability

Variations among studies included in the primary objective were mainly due to study population and setting, assumptions together with model structure. Only 3 of 13 studies synthesised under the primary objective were of the representative population. Apart from deploying different model types, some studies made use of real-time COVID-19 dataset, others used historic datasets while others relied on hypothetical samples. This increased variability and reduced the generalizability of results. However two of three studies implemented in the UK were in favour of the intervention.

An observation of plotted graphs under the secondary objective, showed minimal heterogeneity mostly stemming from the study implemented among pregnant women. Also, a stratification by setting gave a better picture and produced similar rates for studies in the UK and all studies pooled together, besides population level studies.

## Discussion

Albeit considered very low level evidence, review synthesis have shown a clear enough majority vote of 76.9% (95% Binomial Exact CI: 46.2% - 95.0%, P=0.09) in favour of the intervention. Studies that were in favour of the control (Kucharski et al, 2020; Grassly et al, 2020 and Bracis et al, 2020) failed to consider mass testing and contact tracing as a hybrid strategy. These studies went on the assumption that mass testing was not feasible, as acknowledged in an open letter by Peto (2020)(92). Evidence from countries that embarked on mass testing including Taiwan, Germany, Ireland, China and India support the fact that regular mass testing and contact tracing could be the game changer. The analysis by Peto et al (2020) showed that mass testing and contact tracing is by far more cost-effective than present test and trace, in line with the second outcome. Maslov (2020) on the contrary shares an opposing view in that even the slightest false positives will render random mass testing an unreliable policy(93). While Maslov seem to be concerned with the inherent moral decadence of unjust isolation, it is rather better to be on a safe side than in a pool of false negatives and contented asymptomatic carriers. Identification of asymptomatic carriers is crucial because Viswanathan and colleagues also acknowledged that strategies based on symptom screening could miss between 40 – 100% of infected persons(94). Paying attention to asymptomatic infectiousness no matter how small the proportion may be has also been underscored in Byambasuren et al(2020)(95). This argument is also in accordance with the key messages and objectives of European Centre for Disease Prevention and Control, that whole population be tested in high transmission settings(96).

This review also found an overall proportion of asymptomatic carriers among detected positive cases to be 40.7% (95% CI: 38.8-42.5) and 56.6% (95% CI: 49.6 – 63.4) within the UK when stratified. Proportions across studies ranged from 28% among cases detected in the general population to 96.6% among care home staff with positive tests. Also, asymptomatic SARS-CoV-2 prevalence was highest among residents in homeless shelter (30.1%) and lowest among hospital patients (0.0%). The 40.7% asymptomatic proportion among positive cases is in accordance with the 40 – 45% proportion estimated by Oran et al(97). Clarke and colleagues reported a similar rate of 40.3% among haemodialysis patients(98). This proportion is also concordant to that reported in Spain (40.5%) by Albalate et al(99). The proportion of detected positive air travellers (83.6%) found in this review is higher than the 76.6% reported in Al-Qahtani et al(100) perhaps due to more awareness as the study was implemented at a much later date. Yanes-Lane reported an asymptomatic proportion of positive cases among care home residents (54%) just a little lower than the 60.2% reported in this review(101). Notwithstanding the overarching reported high infectivity from asymptomatic individuals, this review reports rates ranging from 0.005 – 1.2% in the population, similar to rates (1.5 – 2%) reported by Wu et al(102). The estimate that the proportion of asymptomatic SARS-CoV-2 among cases in the general population is 28% is in agreement with the community asymptomatic proportion of 28% in Beale et al(103). In contrast, Petersen et al reported a community asymptomatic proportion that was three times higher (76.5% to 86.1%)(104). This population level study was undertaken in the UK, contrary to those included in this review (Iceland, Italy and China). The largest population sample in this review (about a million) was a study done immediately after lockdown which could be the reason behind the low rates.

## Study Limitations

A majority of included studies were modelling studies which normally rely on assumptions that sometimes may hardly be achieved in real life. Expert knowledge was needed to evaluate the validation process of models and it cannot be guarantee that the conclusions on bias were as accurate as would have otherwise expected. The fact that this review went through a single reviewer might have introduced some bias in study selection and analysis. The variability in the understanding of mass testing by different researchers might have had an effect on the analysis as well. This review was language biased since the literature search was limited to English articles. Non peer reviewed articles (preprints) were included in the review thereby reducing the quality of evidence. This review was not registered with PROSPERO for standard systematic review practice and will be erroneous to be considered as such.

## Public Health Implications

Controlling a virus whose manifestation is increasingly without signs is not about number of tests but about who needs to be tested. An appropriate public health strategy that will get the right people tested, at the right time and in the right place requires a community based and participatory approach which will not be without a greater cost burden. Among others, winning the quenched public confidence, ensuring data privacy, acceptability of the NHS app and equity of testing and contact tracing, use of rapid test, capacity building, effective monitoring of isolation and quarantine and programme sustainability are some of the considerations that will have to be made. More real-time research is needed regarding the effectiveness of mass testing and contact tracing, for a better picture of disease burden and mitigation strategies.

## Conclusion and Recommendations

This review sought to critically evaluate the evidence that mass testing and contact tracing is more effective in controlling local transmissions of COVID-19 in the UK, compared to conventional Test and Trace. It has demonstrated a very low level of promising evidence that mass testing and contact tracing could be more effective in bringing the virus under control and even more effective if combined with social distancing and face coverings. The implementation of test and trace has to be done at mass irrespective of symptoms with the local community, through GP surgeries, community health centres and local councils(105). The proposal is for the present Test and Trace to be superseded by a decentralised and continuous mass testing programme with rapid tests, championed by low-resource-need community services(106). The following recommendations are therefore useful; Capacitate GP surgeries and community health services to deliver mass testing at point-of-care.

Government should work in synergy with local councils for robust surveillance, isolation and quarantine(107). This showed major success in Germany(108, 109)

Regular organizational and company-wide testing including the NHS, care homes and schools for the safe return of workforce and students(71, 74).

Coronavirus testing should be a boarder control measure for all travellers(111, 112).

Testing of prisoners, detainees and all those in congested accommodations(55). The Lesbos camp testing in Greece led to more than 240 positive cases(113, 114)

Sewage and environment related testing should be part of mitigation strategies

## Patient Consent

Not applicable

## Declaration of Data Sharing

There is no additional data

## Data Availability

There is no additional data

---

**Supplement 1.** Characteristics of excluded articles

| Study | Design/ Outcome | Sample/ setting | Strategies | Reason for exclusion |
| --- | --- | --- | --- | --- |
| Kucharski(2020)(115)([www.thelancet.com/infection/2020/06/11/S1473-3099(20)30428-2](http://www.thelancet.com/infection/2020/06/11/S1473-3099%2820%2930428-2)) | Modelling study<br>Reduction in transmission<br>Daily contacts quarantined | 40,162 participants and BBC 2017-18 social contact dataset,<br>UK | - No control measure<br>- Self-isolation of symptomatic cases<br>- Household quarantine<br>- Quarantine of work or school contacts<br>- Manual tracing of acquaintances<br>- Manual tracing of all contacts<br>- App-based tracing<br>- Mass testing<br>- Daily limit of other setting contacts | Unsuitable design. No comparison either between intervention and control or between mass testing and symptom-based testing |
| Kirshblum(2020)(116)(https://onlinelibrary.wiley.com/doi/abs/10.1002/pmrj.12454) | Retropective study<br>Test results<br>Symptoms onset | 103 admitted patients in Rehab hospital<br>USA | Analysis of samples collected at time of admission | Unsuitable design |
| Firth et al(117)(https://www.nature.com/articles/s41591-020-1036-8) | Modelling study<br>Number of tests, Number of contacts | 468<br>Real-world social network data,<br>UK | - Outbreak progress under no intervention<br>- Outbreak progress under case isolation<br>- Outbreak progress under primary contact tracing<br>- Outbreak progress under secondary contact tracing | Contact tracing limited to symptom-based testing |
| Keeling et al(118)(https://jech.bmj.com/content/jech/early/2020/06/16/jech-2020-214051.full.pdf) | Cross sectional survey<br>Tracing efficacy<br>Distribution of secondary cases | > 5802 subjects reporting<br>>50,000 contacts<br>UK | N/A | Contact tracing limited to symptom-based testing |
| Bilinski et al(119)(https://jamanetwork.com/journals/jamanetworkopen/fullarticle/2769618?resultClick=1) | Modelling study<br>% reduction in R | Hypothetical,<br>US | - Symptom testing with 30% isolation and quarantine<br>- Test all individuals, with 30% isolation and quarantine<br>- Symptom testing with 60% isolation and quarantine<br>- Test all individuals, with 60% isolation and quarantine<br>-Symptom testing, with 90% isolation and quarantine<br>- Test all individuals, with 90% isolation and quarantine | Contact tracing limited to symptom-based testing |
| Kretzschmar et al(120)([https://www.thelancet.com/journals/lanpub/article/PIIS2468-2667(20)30157-2/fulltext](https://www.thelancet.com/journals/lanpub/article/PIIS2468-2667%2820%2930157-2/fulltext)) | Modelling study<br>Reduction in Reproduction number | Hypothetical,<br>Netherlands | - Conventional contact tracing<br>- Mobile app contact tracing<br>- Physical distancing strategy<br>- Testing and isolation of cases without tracing contacts | Contact tracing limited to symptom-based testing |
| Skoll(2020)(121)(https://www.sciencedirect.com/s) | Non-systematic review<br>- Role of technology | N/A | - Digital contact tracing and mass testing | Unsuitable design |
| cience/article/pii/S2666693620300360) | - Barriers<br>- Scale-up strategies |  |  |  |
| Kerr(2020)(122)(http://medrxiv.org/content/early/2020/07/16/2020.07.15.20154765.abstract) | Modelling study<br>Feasibility of control strategies | Demographic, mobility and epidemiological data of Seattle USA | Test and trace (testing, contact tracing and quarantine) | Limited to control |
| Panovska-Griffiths(2020)(123)(https://www.thelancet.com/action/showPdf?pii=S2352-4642%2820%2930250-9) | Modelling study<br>Reduction in Reproduction number | Modelled sample, UK | - Full time schooling<br>- Part-time weekly rota system of 50% each schooling<br>- 68% contact tracing with no scale-up in testing<br>- 68% contact tracing with sufficient testing<br>- 40% contact tracing with sufficient testing | No suitable comparison |
| Hellewell(2020)(124)([https://www.thelancet.com/journals/langlo/article/PIIS2214-109X(20)30074-7/fulltext](https://www.thelancet.com/journals/langlo/article/PIIS2214-109X%2820%2930074-7/fulltext)) | Modelling study<br>Onward transmission | Modelled sample, UK | - 5, 20 and 40 initial cases of outbreak<br>- 0, 0.2, 0.4, 0.6, 0.8 and 1 probabilities of tracing a contact<br>- Short symptom onset to isolation<br>- Long symptom onset to isolation | Contact tracing limited to symptom-based testing |
| Ferretti(2020)(125)(https://www.ncbi.nlm.nih.gov/pmc/articles/PMC7164555/) | Modelling study<br>Basic reproduction number – R Generation time | Pair of 40 hypothetical recipients<br>Singapore | - Symptomatic transmission<br>- Presymptomatic transmission<br>- Asymptomatic transmission<br>- Environmental transmission<br>- Isolating symptomatic persons<br>- Tracing the contacts of symptomatic cases and quarantining | Contact tracing limited to symptom-based testing |
| Min(2020)(126)(http://www.ncbi.nlm.nih.gov/pubmed/32893522) | Modelling study<br>Epidemic size<br>Effective contact rate | Daily reported cases (Feb12-March3)<br>Korea | - Social distancing among adults<br>- Spring semester postponement<br>- Intensive contact tracing<br>- Large-scale diagnostic testing | No suitable comparator |
| Quilty(2020)(111)(https://www.eurosurveillance.org/docserver/fulltext/eurosurveillance/25/5/eurosurv-25-5-2.pdf) | Modeling study<br>Infected travellers | Air travellers | - Symptoms screening | Unsuitable design |
| Domenico(2020)(106)(https://bmcmecicine.biomedcentral.com/articles/10.1186/s12916-020-01698-4) | Modelling study<br>- Lockdown impact<br>- Number of contacts | Age profile data of Ile-de-France and 2012 social contact matrix | - School closure<br>- Employee telework at from home<br>- Senior isolation (high risk group)<br>- Lockdown and non-essential activity ban<br>- Case isolation with large-scale testing | Unsuitable comparator |
| Gostic(2020)(112)(https://elifesciences.org/article/s/55570) | Modelling study<br>Screening outcome and missed cases | Hypothetical population of infected travellers, | - Symptomatic but not aware of exposure risk<br>- Aware of exposure risk but without detectable symptoms<br>- Symptomatic and aware that exposure may have occurred<br>- Neither symptomatic nor aware of exposure risk | Lack of intervention |
| He(2020)(127)(https://rs-delve.github.io/pdfs/2020-05-27-effectiveness-and-resource-requirements-of-tti-strategies.pdf) | Modelling study<br>Resources required<br>Effect on R | 40,162 BBC pandemic data<br>UK | - Symptom-based contact tracing<br>- Test-based contact tracing<br>- Testing of asymptomatic contacts | Contact tracing limited to symptom-based testing |
| Goscé(2020)(128) (https://www.sciencedirect.com/science/article/pii/S0163445320303157?pes=vor) | Modelling study | PHE, NHS and TfL data<sup>5</sup><br>UK<br>Royal Borough of Kensington and Chelsea (RBKC) | - Isolation of RBKC residents from the rest of the city<br>- Removal of lockdown<br>- Weekly testing (business reopens but people work from home)<br>- Shielding 60+ age group with lifting of lockdown<br>- Combined universal testing and use of face coverings with no lockdown<br>- Universal testing, contact tracing and isolation, lockdown | Unsuitable design |
| Li(2020)(19)(https://www.sciencedirect.com/science/article/pii/S0163445320303157?pes=vor) | Descriptive study | N/A | - Containment<br>- Suppression | Unsuitable design |
| Kennedy-Shaffer(2020)(129)(https://dash.harvard.edu/handle/1/37363184) | Modelling study<br>Reduction in transmissions | Unknown | - Hypothetical rapid test<br>- Transmission tracing<br>- Full isolation of all contacts of cases<br>- Isolate contacts with positive test result | Unsuitable design |
| Maslov(2020)((93)(https://papers.ssrn.com/abstract=3643408) | Logical description<br>Economic benefits | Non | Random mass testing | No comparator |
| Peto(2020)(130) (https://royalsocietypublishing.org/doi/pdf/10.1098/rsos.200915) | Modelling study<br>Reproduction number<br>Number of daily tests | Hypothetical<br>UK | - Weekly mass test and trace using isothermal single-step RT-PCR | No comparator |
| Campbell(2020)(131)(https://www.cmaj.ca/content/early/2020/09/09/cmaj.201128) | Cross sectional<br>Cost, human resource and lab capacity | 41 751<br>COVID-19 contacts, staff of hospitals, health centres, care homes & | - Systematic trace and test contacts<br>- Test all staff of acute care hospitals<br>- Test all community health workers and staff/residents of long-term care homes | No suitable comparator |

<sup>5</sup> PHE : Public Health England, NHS : National Health Service, TfL : Transport for London

| Study | Design/ Outcome | Sample/ setting | Strategies | Reason for exclusion |
| --- | --- | --- | --- | --- |
|  |  | essential businesses,<br>school children & staff<br>Canada | - Test all major public and interpersonal contact essential workers<br>- Test all children and staff of schools |  |
| Cleevely(2020)(132)(http://academic.oup.com/oxrep/article/36/Supplement_1/S14/5899015) | Modelling study | Hypothetical<br>UK | - Stratified periodic sample testing<br>- Universal random testing | Unsuitable comparator |
| Yokota(2020)(133)(https://www.ncbi.nlm.nih.gov/pmc/articles/PMC7543374/) | Diagnostic tests<br>Utility of nucleic acid amplification | 1,924 asymptomatic persons<br>Japan | - Nasopharyngeal swap-based (NPS) RT-PCR test<br>- Saliva-based PCR test | Unsuitable design |
| Eilersen(2020)(134)(https://doi.org/10.1038/s41598-020-75640-2) | Modelling study<br>Quarantine measures<br>Cost-effectiveness | Hypothetical | - No intervention<br>- Reduced work contacts by 75%<br>- Reduced social contacts by 75%<br>- Infection probability reduced by 50%<br>- Workplace size reduced by half<br>- Infection probability plus workplace size reduced | Limited to control |
| Altawalah(2020)(135)(https://www.ncbi.nlm.nih.gov/pmc/articles/PMC7527795/) | Cross sectional study | 891 suspects<br>Kuwait | - Nasopharyngeal swap-based (NPS) RT-PCR test<br>- Saliva-based PCR test | Unsuitable design |
| Dollard(2020)(136)(http://www.cdc.gov/mmwr/volumes/69/wr/mm6945a4.htm?s_cid=mm6945a4_w) | Diagnostic test<br>COVID-19 infections | 298 air travellers<br>USA | RT-PCR | Asymptomatic proportions unknown |
| Telford(2020)(137)(https://gov.georgia.gov/executive-action/e) | Cross sectional<br>Timing of mass testing | 5,671 residents & staff in 28 long term care facility<br>USA | Mass RT-PCR test | Asymptomatic proportion unknown |
| Bosetti(2020)(138)(https://doi.org/10.1101/2020.12.08.20246009) | Modelling study<br>Impact of intervention | Real-time data<br>France | Mass testing | No comparator |

**Supplement 2.**
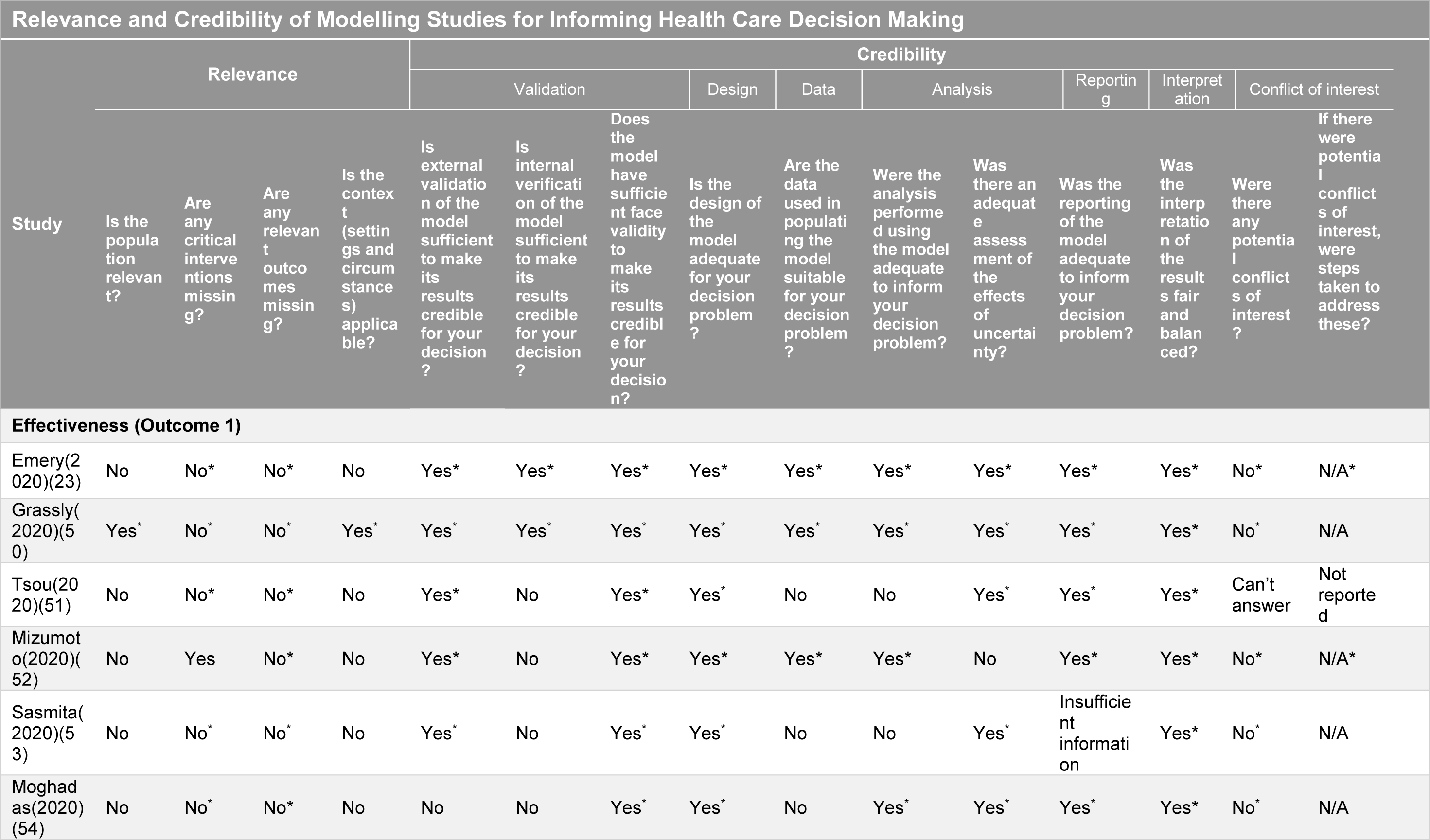
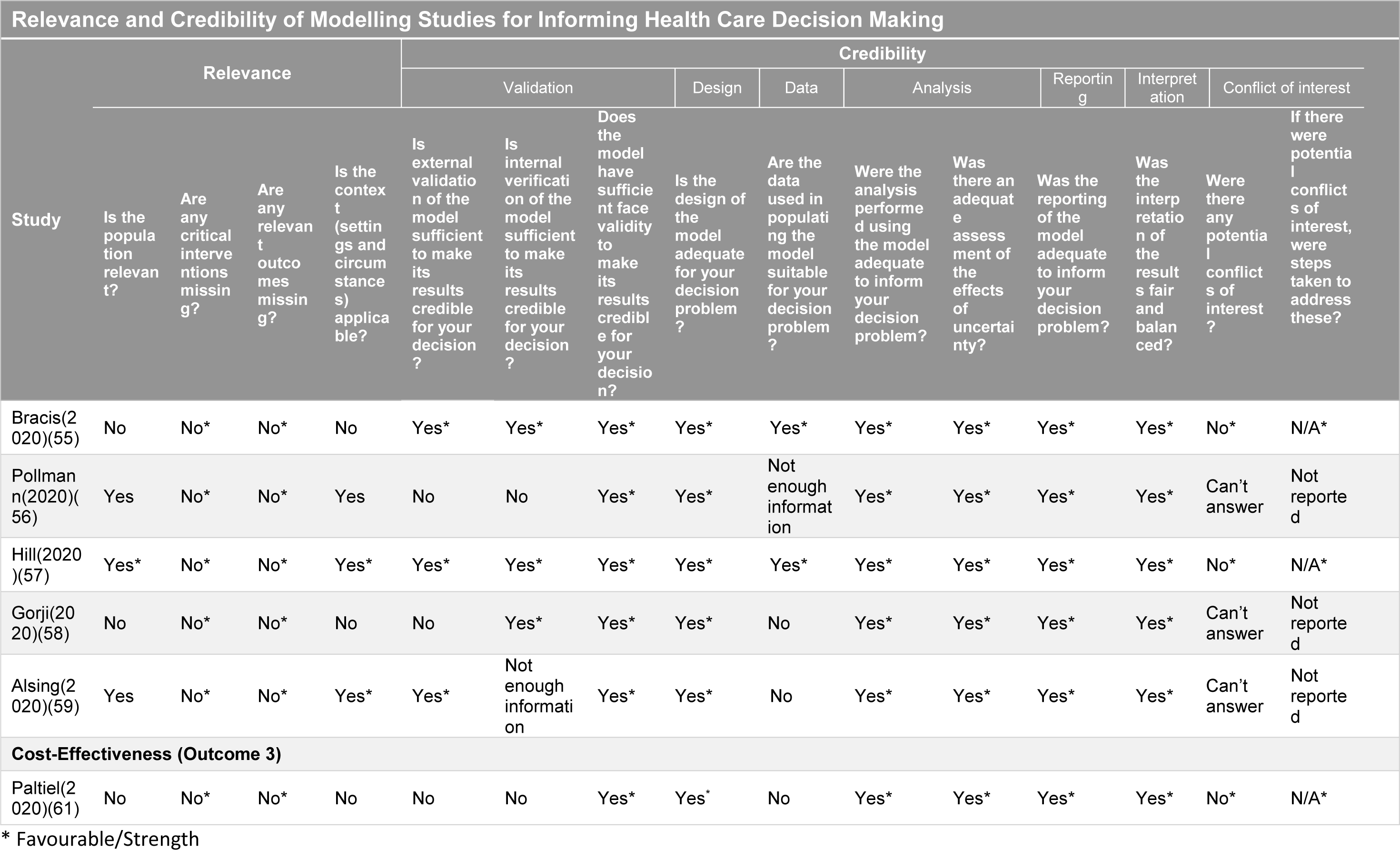
Methodological quality assessment of modelling studies

**Supplement 3.** Methodological quality assessment of Cohort Studies

| Critical Appraisal Skills Programme (CASP) Tool |  |  |  |  |  |  |  |  |  |  |  |  |  |  |
| --- | --- | --- | --- | --- | --- | --- | --- | --- | --- | --- | --- | --- | --- | --- |
| Are the results of the study valid? |  |  |  |  |  |  |  |  | What are the results? |  |  |  |  |  |
| Study | Did the study address a clearly focused issue? | Was the cohort recruited in an acceptable way? | Was the exposure accurately measured to minimise bias? | Was the outcome accurately measured to minimise bias? | Have the authors identified all important confounding factors? | Have they taken account of the confounding factors in the design and/or analysis? | Was the follow up of subjects complete enough? | Was the follow up of subjects long enough? | What are the results of this study?<sup>4</sup> | How precise are the results? | Do you believe the results? | Can the results be applied to the local population? | Do the results of this study fit with other available evidence? | What are the implications of this study for practice? |
| **Effectiveness (Outcome 1)** |  |  |  |  |  |  |  |  |  |  |  |  |  |  |
| Porru(20)(63)(<br>https://www.mdpi.com/1660-4601/17/14/5104) | Yes | Yes | Yes | Yes<sup>1</sup> | No<sup>2</sup> | Yes | Yes<sup>3</sup> | Yes | A total 238 workers had a positive test with a cumulative incidence of 4.0%. A third of positive cases were symptomatic while 2.3% were asymptomatic; Incidence among the exposed was 40.5% and 0.5% among the non-exposed group | Cumulative incidence was 4.0% (95% CI 3.5–4.5%). The risk odds ratio in medical wards was 2.7 (95% CI 1.9–3.9) and 4.3 (95% CI 2.4–7.6) in health services | Yes | No<sup>5</sup> | Yes | Yes<sup>5</sup> |
| Comments<br>1. Neither subjects nor outcome assessors were blinded<br>2. Ethnicity of healthcare workers might have been important<br>3. No report on whether there was loss to follow-up or not<br>4. Absolute risk reduction unreported<br>5. Limited applicability due to study population and contextual differences<br>6. Study highlights the importance of mass testing and contact tracing in a healthcare setting which can be applicable to any context |  |  |  |  |  |  |  |  |  |  |  |  |  |  |

**Supplement 5.**
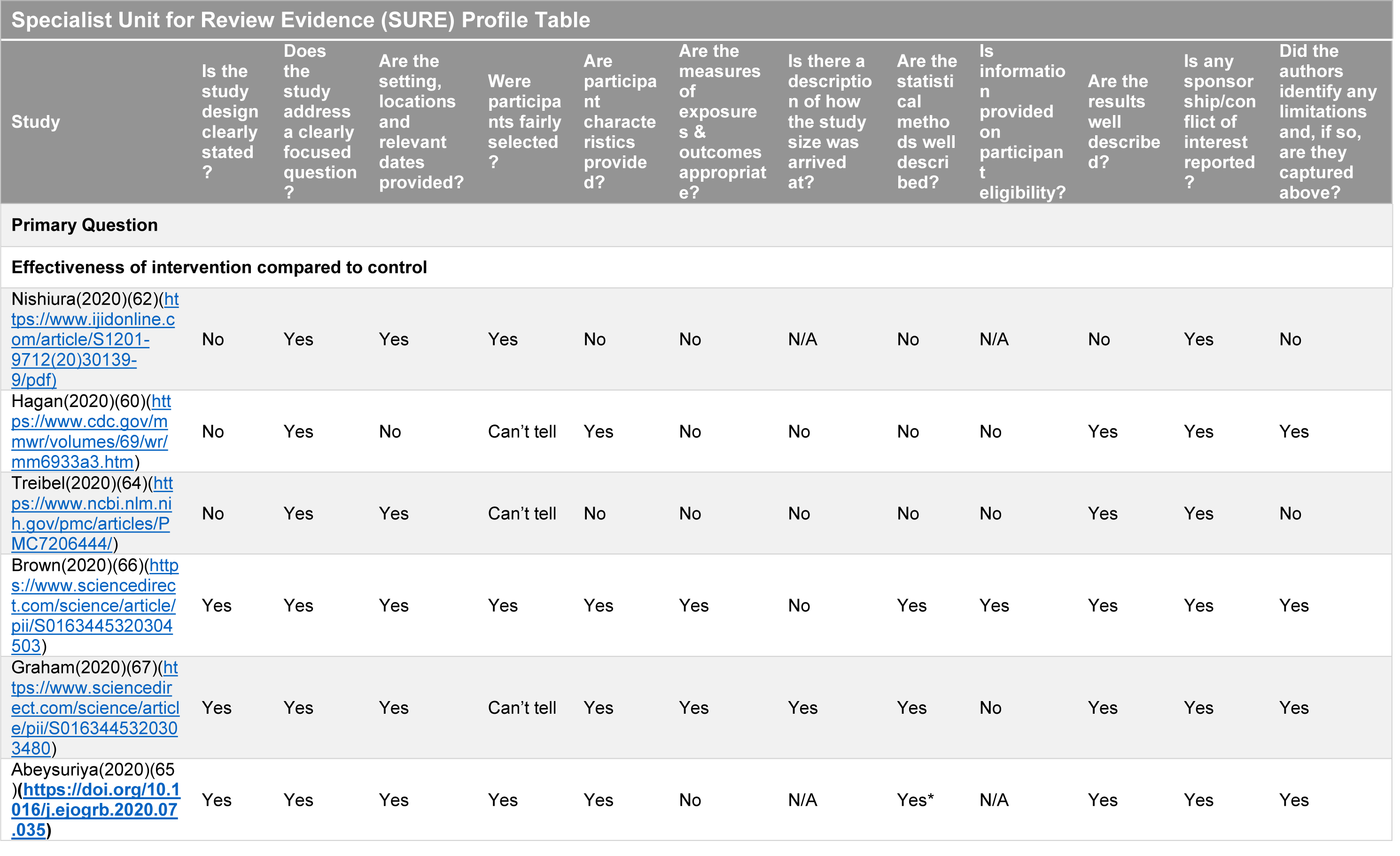
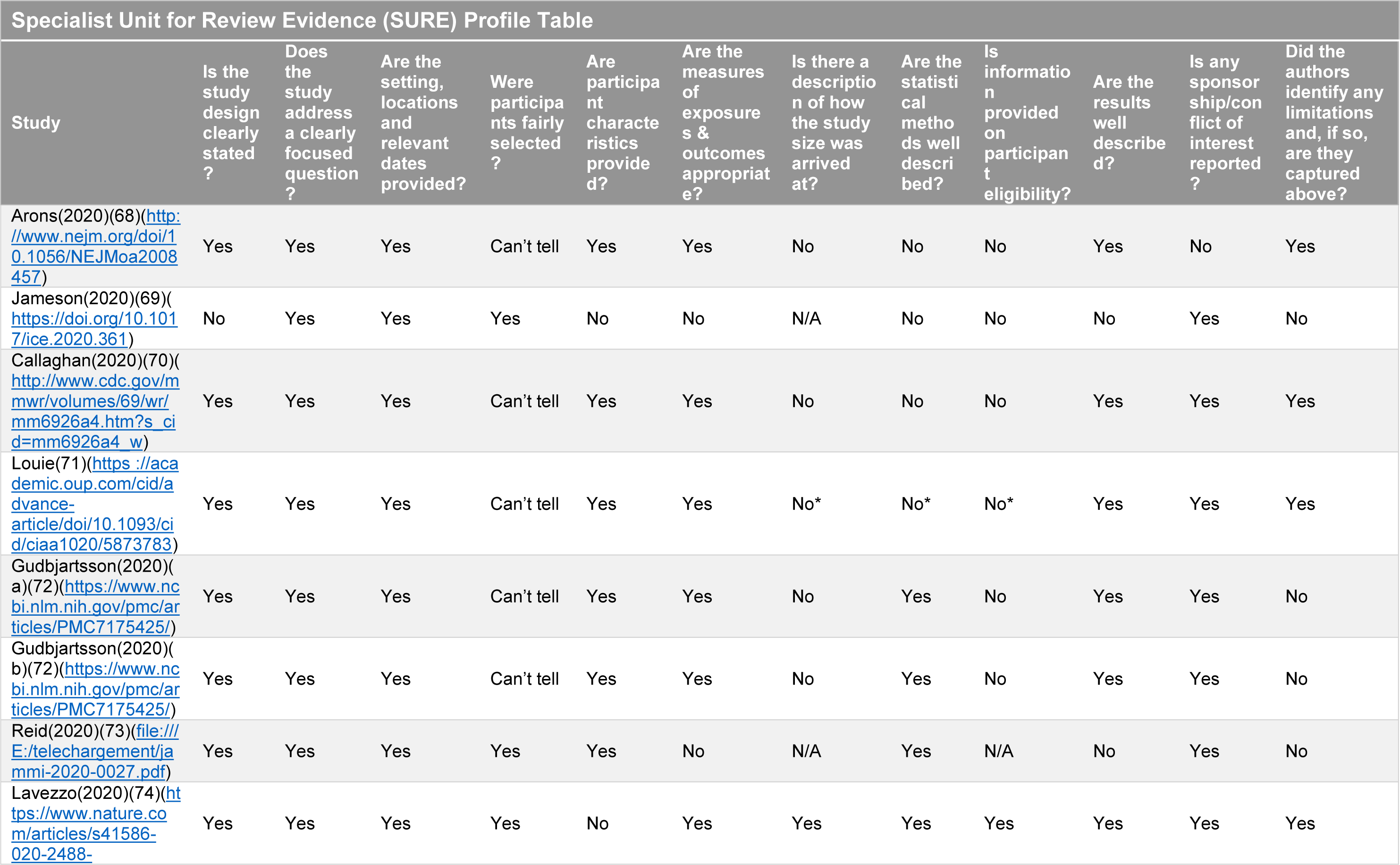
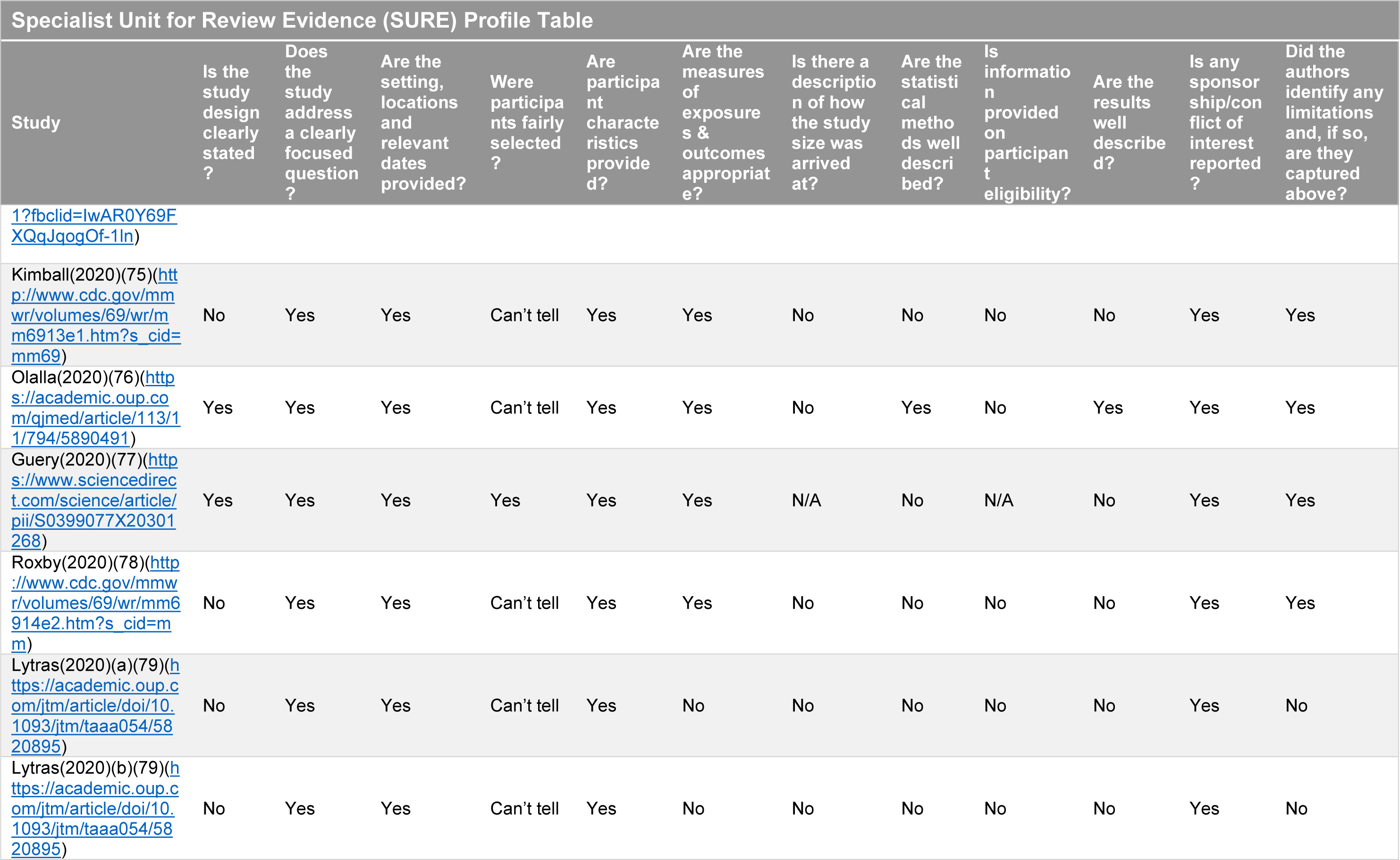
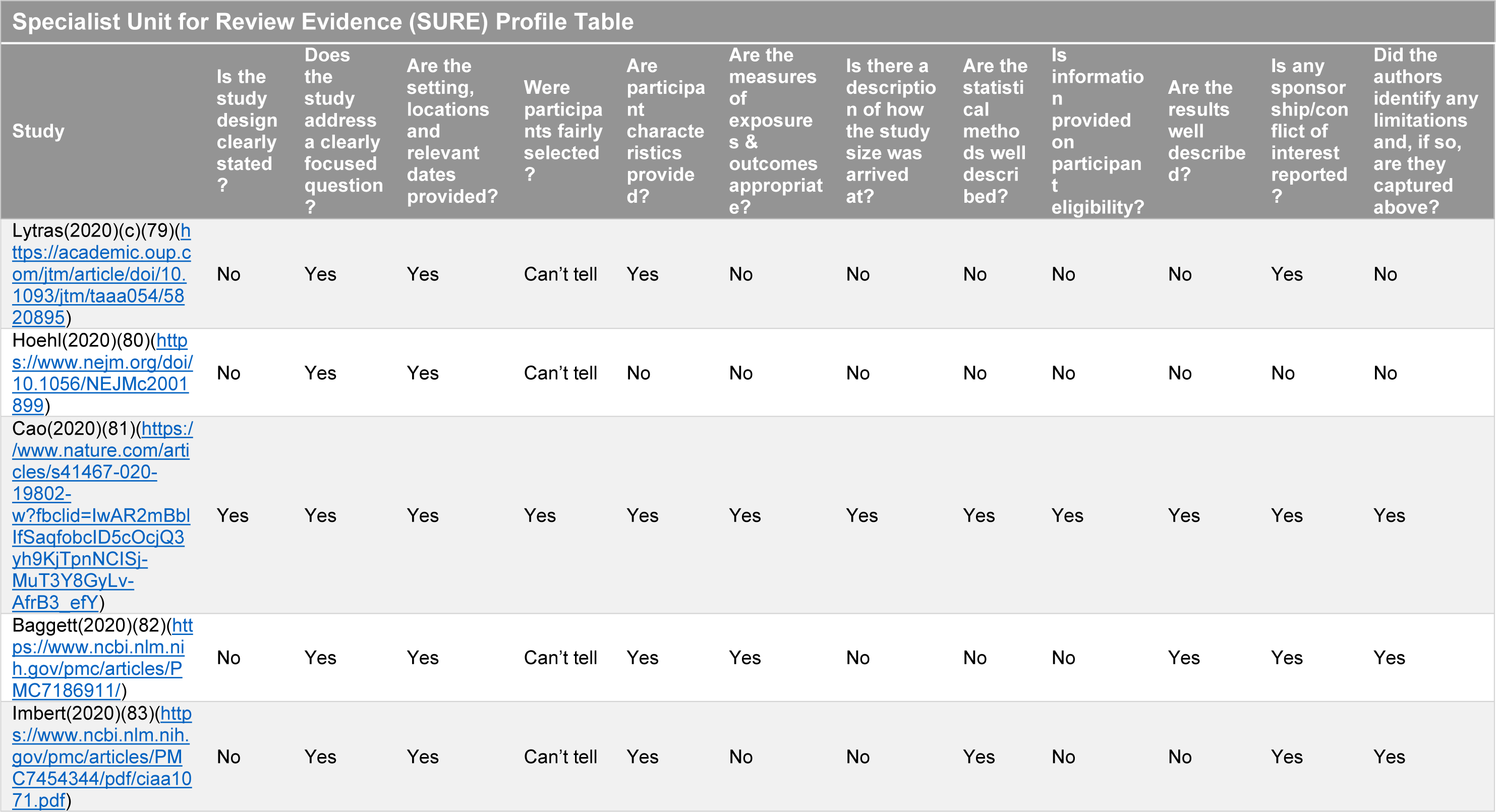
Methodological quality assessment of cross sectional studies

| Specialist Unit for Review Evidence (SURE) Profile Table |  |  |  |  |  |  |  |  |  |  |  |  |
| --- | --- | --- | --- | --- | --- | --- | --- | --- | --- | --- | --- | --- |
| Study | Is the study design clearly stated? | Does the study address a clearly focused question? | Are the setting, locations and relevant dates provided? | Were participants fairly selected? | Are participant characteristics provided? | Are the measures of exposures & outcomes appropriate? | Is there a description of how the study size was arrived at? | Are the statistical methods well described? | Is information provided on participant eligibility? | Are the results well described? | Is any sponsorship/conflict of interest reported? | Did the authors identify any limitations and, if so, are they captured above? |
| **Primary Question** |  |  |  |  |  |  |  |  |  |  |  |  |
| **Effectiveness of intervention compared to control** |  |  |  |  |  |  |  |  |  |  |  |  |
| Nishiura(2020)(62)([https://www.ijidonline.com/article/S1201-9712(20)30139-9/pdf](https://www.ijidonline.com/article/S1201-9712%2820%2930139-9/pdf)) | No | Yes | Yes | Yes | No | No | N/A | No | N/A | No | Yes | No |
| Hagan(2020)(60)(https://www.cdc.gov/mmwr/volumes/69/wr/mm6933a3.htm) | No | Yes | No | Can't tell | Yes | No | No | No | No | Yes | Yes | Yes |
| Treibel(2020)(64)(https://www.ncbi.nlm.nih.gov/pmc/articles/PMC7206444/) | No | Yes | Yes | Can't tell | No | No | No | No | No | Yes | Yes | No |
| Brown(2020)(66)(https://www.sciencedirect.com/science/article/pii/S0163445320304503) | Yes | Yes | Yes | Yes | Yes | Yes | No | Yes | Yes | Yes | Yes | Yes |
| Graham(2020)(67)(https://www.sciencedirect.com/science/article/pii/S0163445320303480) | Yes | Yes | Yes | Can't tell | Yes | Yes | Yes | Yes | No | Yes | Yes | Yes |
| Abeyasuriya(2020)(65)(https://doi.org/10.1016/j.ejogrb.2020.07.035) | Yes | Yes | Yes | Yes | Yes | No | N/A | Yes* | N/A | Yes | Yes | Yes |
| Study | Is the study design clearly stated? | Does the study address a clearly focused question? | Are the setting, locations and relevant dates provided? | Were participants fairly selected? | Are participant characteristics provided? | Are the measures of exposure & outcomes appropriate? | Is there a description of how the study size was arrived at? | Are the statistical methods well described? | Is information provided on participant eligibility? | Are the results well described? | Is any sponsorship/conflict of interest reported? | Did the authors identify any limitations and, if so, are they captured above? |
| Arons(2020)(68)(http://www.nejm.org/doi/10.1056/NEJMoa2008457) | Yes | Yes | Yes | Can't tell | Yes | Yes | No | No | No | Yes | No | Yes |
| Jameson(2020)(69)(https://doi.org/10.1017/ice.2020.361) | No | Yes | Yes | Yes | No | No | N/A | No | No | No | Yes | No |
| Callaghan(2020)(70)(http://www.cdc.gov/mmwr/volumes/69/wr/mm6926a4.htm?s_cid=mm6926a4_w) | Yes | Yes | Yes | Can't tell | Yes | Yes | No | No | No | Yes | Yes | Yes |
| Louie(71)(https://academic.oup.com/cid/article/doi/10.1093/cid/ciaa1020/5873783) | Yes | Yes | Yes | Can't tell | Yes | Yes | No* | No* | No* | Yes | Yes | Yes |
| Gudbjartsson(2020)(a)(72)(https://www.ncbi.nlm.nih.gov/pmc/articles/PMC7175425/) | Yes | Yes | Yes | Can't tell | Yes | Yes | No | Yes | No | Yes | Yes | No |
| Gudbjartsson(2020)(b)(72)(https://www.ncbi.nlm.nih.gov/pmc/articles/PMC7175425/) | Yes | Yes | Yes | Can't tell | Yes | Yes | No | Yes | No | Yes | Yes | No |
| Reid(2020)(73)(file:///E:/telechargement/jammi-2020-0027.pdf) | Yes | Yes | Yes | Yes | Yes | No | N/A | Yes | N/A | No | Yes | No |
| Lavezzo(2020)(74)(https://www.nature.com/articles/s41586-020-2488- | Yes | Yes | Yes | Yes | No | Yes | Yes | Yes | Yes | Yes | Yes | Yes |

Specialist Unit for Review Evidence (SURE) Profile Table
| Study | Is the study design clearly stated? | Does the study address a clearly focused question? | Are the setting, locations and relevant dates provided? | Were participants fairly selected? | Are participant characteristics provided? | Are the measures of exposure & outcomes appropriate? | Is there a description of how the study size was arrived at? | Are the statistical methods well described? | Is information provided on participant eligibility? | Are the results well described? | Is any sponsorship/conflict of interest reported? | Did the authors identify any limitations and, if so, are they captured above? |
| --- | --- | --- | --- | --- | --- | --- | --- | --- | --- | --- | --- | --- |
| 1?fbclid=IwAR0Y69FXQqJqogOf-1ln) |  |  |  |  |  |  |  |  |  |  |  |  |
| Kimball(2020)(75)(http://www.cdc.gov/mmwr/volumes/69/wr/mm6913e1.htm?s_cid=mm69) | No | Yes | Yes | Can't tell | Yes | Yes | No | No | No | No | Yes | Yes |
| Olalla(2020)(76)(http://academic.oup.com/qjmed/article/113/11/794/5890491) | Yes | Yes | Yes | Can't tell | Yes | Yes | No | Yes | No | Yes | Yes | Yes |
| Guery(2020)(77)(http://www.sciencedirect.com/science/article/pii/S0399077X20301268) | Yes | Yes | Yes | Yes | Yes | Yes | N/A | No | N/A | No | Yes | Yes |
| Roxby(2020)(78)(http://www.cdc.gov/mmwr/volumes/69/wr/mm6914e2.htm?s_cid=mm6914e2) | No | Yes | Yes | Can't tell | Yes | Yes | No | No | No | No | Yes | Yes |
| Lytras(2020)(a)(79)(https://academic.oup.com/jtm/article/doi/10.1093/jtm/taaa054/5820895) | No | Yes | Yes | Can't tell | Yes | No | No | No | No | No | Yes | No |
| Lytras(2020)(b)(79)(https://academic.oup.com/jtm/article/doi/10.1093/jtm/taaa054/5820895) | No | Yes | Yes | Can't tell | Yes | No | No | No | No | No | Yes | No |
| Study | Is the study design clearly stated? | Does the study address a clearly focused question? | Are the setting, locations and relevant dates provided? | Were participants fairly selected? | Are participant characteristics provided? | Are the measures of exposure & outcomes appropriate? | Is there a description of how the study size was arrived at? | Are the statistical methods well described? | Is information provided on participant eligibility? | Are the results well described? | Is any sponsorship/conflict of interest reported? | Did the authors identify any limitations and, if so, are they captured above? |
| Lytras(2020)(c)(79)(https://academic.oup.com/jtm/article/doi/10.1093/jtm/taaa054/5820895) | No | Yes | Yes | Can't tell | Yes | No | No | No | No | No | Yes | No |
| Hoehl(2020)(80)(https://www.nejm.org/doi/10.1056/NEJMc2001899) | No | Yes | Yes | Can't tell | No | No | No | No | No | No | No | No |
| Cao(2020)(81)(https://www.nature.com/articles/s41467-020-19802-w?fbclid=IwAR2mBblfSagfobclD5cOcjQ3yh9KjTpnNCISj-MuT3Y8GyLv-AfrB3_efY) | Yes | Yes | Yes | Yes | Yes | Yes | Yes | Yes | Yes | Yes | Yes | Yes |
| Baggett(2020)(82)(https://www.ncbi.nlm.nih.gov/pmc/articles/PMC7186911/) | No | Yes | Yes | Can't tell | Yes | Yes | No | No | No | Yes | Yes | Yes |
| Imbert(2020)(83)(https://www.ncbi.nlm.nih.gov/pmc/articles/PMC7454344/pdf/ciaa1071.pdf) | No | Yes | Yes | Can't tell | Yes | No | No | Yes | No | No | Yes | Yes |

**Supplement 6.**
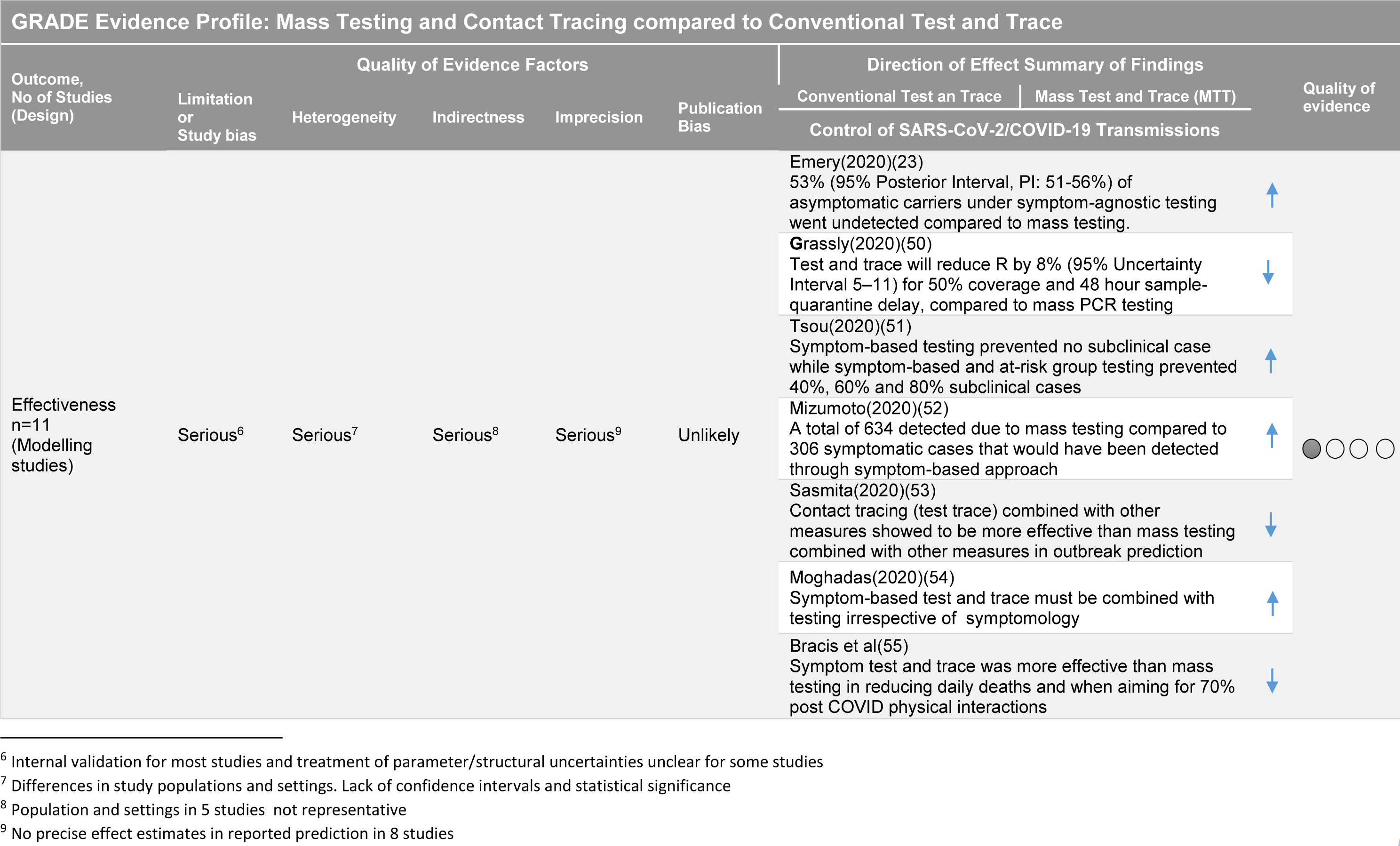
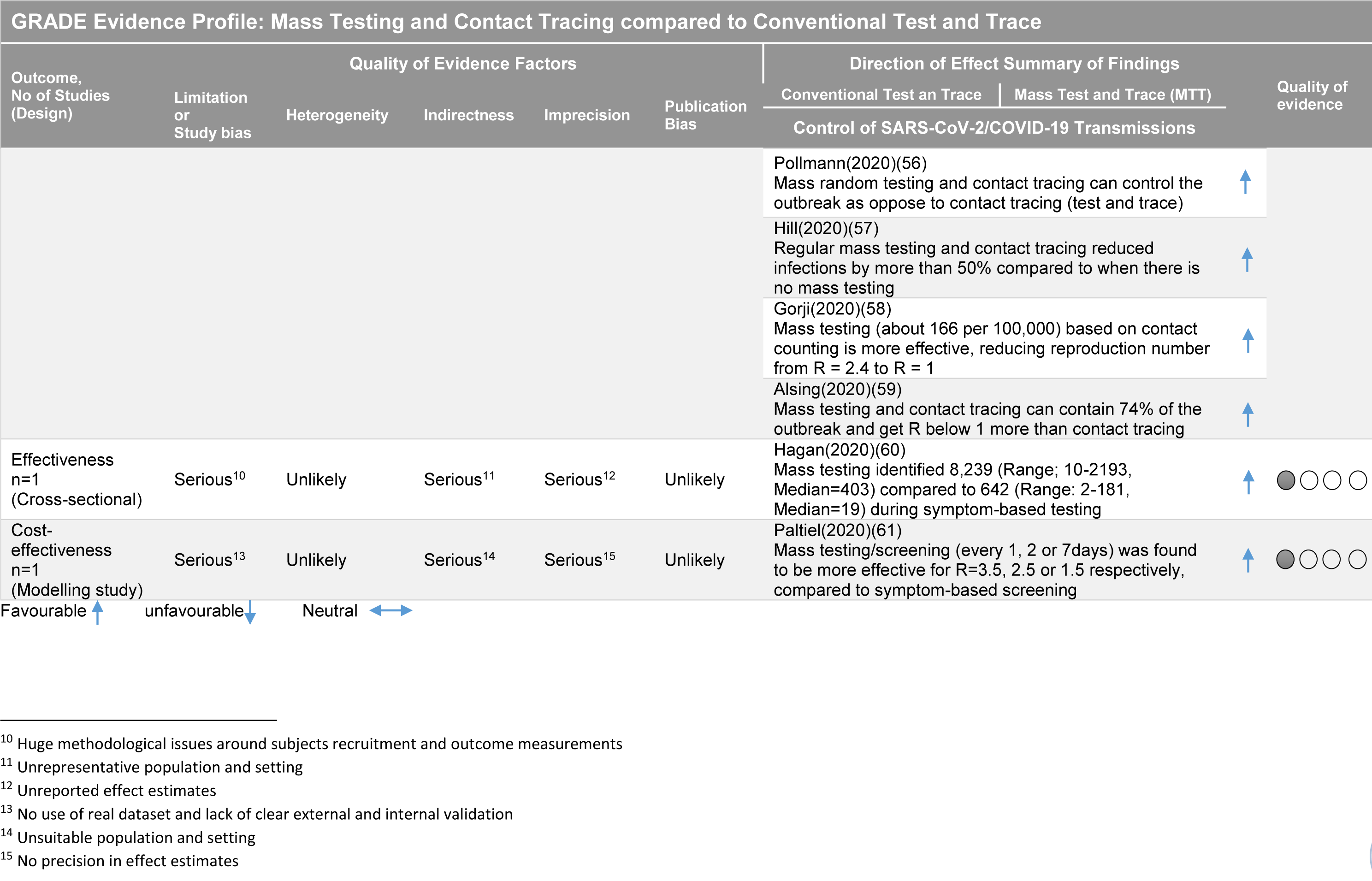
Certainty of Evidence for the Primary Objective

**Supplement 7.** Proportion of detected asymptomatic cases during mass testing

| Study | Test Details | Cases | Asymptomatic | Sample | % in Cases | (95% CI) | % in Sample | (95% CI) |
| --- | --- | --- | --- | --- | --- | --- | --- | --- |
| Secondary Question |  |  |  |  |  |  |  |  |
| Proportion of Asymptomatic (Outcome 6) |  |  |  |  |  |  |  |  |
| Nishiura(2020)(62) | Reverse transcription PCR | 13 | 4 | 565 | 30.8% | (7.7– 53.8%) | 0.7% | (0.2 – 1.8%) |
| Porru(2020)(63) | real-time PCR (Seegene AllplexTM2019-nCoV Assay) on oropharyngeal and nasopharyngeal swabs | 238 | 109 | 5942 | 45.8% | (39.3 – 52.4%) | 1.8% | (1.5 – 2.2%) |
| Treibel(2020)(64)(staffw1) |  | 28 | 28 | 396 | 100% | (87.6 – 100%) | 7.1% | (4.9%–10.0%) |
| Treibel(2020)(64)(staffw2) |  | 14 | 14 | 284 | 100% | (76.8 – 100%) | 4.9% | (3.0%–8.1%) |
| Treibel(2020)(64)(staffw3) |  | 4 | 4 | 263 | 100% | (39.8 – 100%) | 1.5% | (0.6%–3.8%) |
| Treibel(2020)(64)(staffw4) |  | 4 | 4 | 267 | 100% | (39.8 – 100%) | 1.5% | (0.6%–3.8%) |
| Treibel(2020)(64)(staffw5) |  | 3 | 3 | 269 | 100% | (29.2 – 100%) | 1.1% | (0.4%–3.2%) |
| Brown(2020)(66) | Applied Biosystems 7500 FAST system and ELITE InGenius (OSANG Healthcare) | 23 | 4 | 1152 | 17.4% | (5.0 – 38.8%) | 0.3% | (0.1 – 0.9%) |
| Graham(2020)(67)(redts) | AusDiagnostics, Roche Cobas and Abbott RealTime SARS- CoV-2 assays | 126 | 54 | 313 | 42.9% | (34%–52%). | 17.3% | (13.2 – 21.9%) |
| Graham(2020)(67)(staff) |  | 3 | 3 | 70 | 100% | (29.2 – 100%) | 4.3% | (1% to 9%) |
| Abeyasuriya(2020)(65) |  | 7 | 6 | 180 | 85.7% | (42.1–99.6%) | 3.3% | (1.2 – 7.1%) |
| Arons(2020)(68) | SARS-CoV-2 CDC assay protocol | 48 | 27 | 76 | 56.3% | (41.2 – 70.5%) | 35.5% | (24.9 – 47.3%) |
| Jameson(2020)(69) | GeneXpert RT-PCR assay (Cepheid, Sunnyvale, CA) | 0 | 0 | 121 | - | - | 0.0% | (0.0 – 1.2%) |
| Callaghan(2020)(70) | (RT)–PCR Diagnostic Panel | 0 | 0 | 247 | - | - | 0.0% | (0.0 – 1.2%) |
| Louie(71)(redts-staff) | Abbott m2000 RT-PCR | 86 | 86 | 303 | 100% | (95.8 – 100%) | 28.4% | (23.4 – 33.8%) |
| Gudbjartsson(2020)(72)(targettest1) | TaqMan™ Fast Virus 1- step Master Mix, 2019-nCoV Assay kits v1 (Thermo Fisher) | 177 | 24 | 1924 | 13.6% | (8.9 – 19.5%) | 1.2% | (0.8 – 1.9%) |
| Gudbjartsson(2020)(72)(poptest) |  | 87 | 51 | 10797 | 58.6% | (47.6 – 69.1%) | 0.5% | (0.4 – 0.6%) |
| Gudbjartsson(2020)(72)(targettest2) |  | 1044 | 59 | 7275 | 5.7% | (4.3 – 7.2%) | 0.8% | (0.6 – 1.0%) |
| Gudbjartsson(2020)(72)(randomtest1) |  | 13 | 7 | 2283 | 53.8% | (25.1 – 80.8%) | 0.3% | (0.1 – 0.6%) |
| Reid(2020)(73) |  | 5 | 5 | 2751 | 100% | (47.8 – 100%) | 0.2% | (0.1 – 0.2%) |
| Lavezzo(2020)(74)(survey1) | TaqMan Fast Virus 1-Step Master Mix (Thermo Fisher) | 73 | 29 | 2812 | 39.7% | (28.5–51.9) | 1.0% | (0.7 – 1.5%) |
| Lavezzo(2020)(74)(survey2) |  | 29 | 13 | 2343 | 44.8% | (26.5–64.3) | 0.6% | (0.3 – 0.9%) |
| Kimball(2020)(75) | SARS-CoV-2 CDC assay protocol | 23 | 13 | 76 | 56.5% | (34.5 – 76.8%) | 17.1% | (9.4 – 27.5%) |
| Olalla(2020)(76) | VIASURE SARS-CoV-2 from CerTest Biotec and LightMix Modular SARS-CoV (COVID19), Roche | 2 | 1 | 498 | 50.0% | (1.3 – 98.7%) | 0.2% | (0 – 1.1%) |
| Guery(2020)(77) | Detection kit for 2019 novel coronavirus RNA, PCR Fluorescence Probing, (Daan Gene Co.) | 3 | 2 | 136 | 66.7% | (9.4 – 99.2%) | 1.5% | (0.2 – 5.2%) |
| Roxby(2020)(78)(re dts-staffd1) |  | 6 | 3 | 142 | 40.0% | (9.4 – 99.2%) | 2.1% | (0.4 – 6.0%) |
| Lytras(2020)(79)(UK) |  | 13 | 13 | 357 | 100% | (75.3 – 100%) | 3.6% | (2.0%–6.1%) |
| Lytras(2020)(79)(Spain) |  | 25 | 25 | 394 | 100% | (86.3 – 100%) | 6.3% | (4.1%–9.2%) |
| Lytras(2020)(79)(Turkey) |  | 2 | 2 | 32 | 100% | (15.8 – 100%) | 6.3% | (0.8%–20.8%) |
| Hoehl(2020)(80) | LightMix Modular SARS and Wuhan CoV E-gene, and LightMix Modular Wuhan CoV RdRP-gen (TIB MOLBIOL) | 2 | 2 | 114 | 100% | (15.8 – 100%) | 1.8% | (0.2 – 6.2%) |
| Cao(2020)(81) | Real-time fluorescence RT-PCR | 300 | 300 | 9899828 | 100% | (98.8 – 100%) | 0.0% | 0% |
| Baggett(2020)(82) |  | 147 | 129 | 408 | 87.8% | (81.3 – 92.6%) | 31.6% | (27.1 – 36.4%) |
| Imbert(2020)(83) | Abbott m2000 RT-PCR | 111 | 57 | 210 | 51.4% | (41.7 – 61%) | 27.1% | (21.3 – 33.1%) |
| **TOTAL** |  | **2659** | **1081** | **9942968** | **40.65%** | **(38.8 – 42.5%)** | **0.01%** | **(0.01 – 0.012)** |

**Supplement 8.**
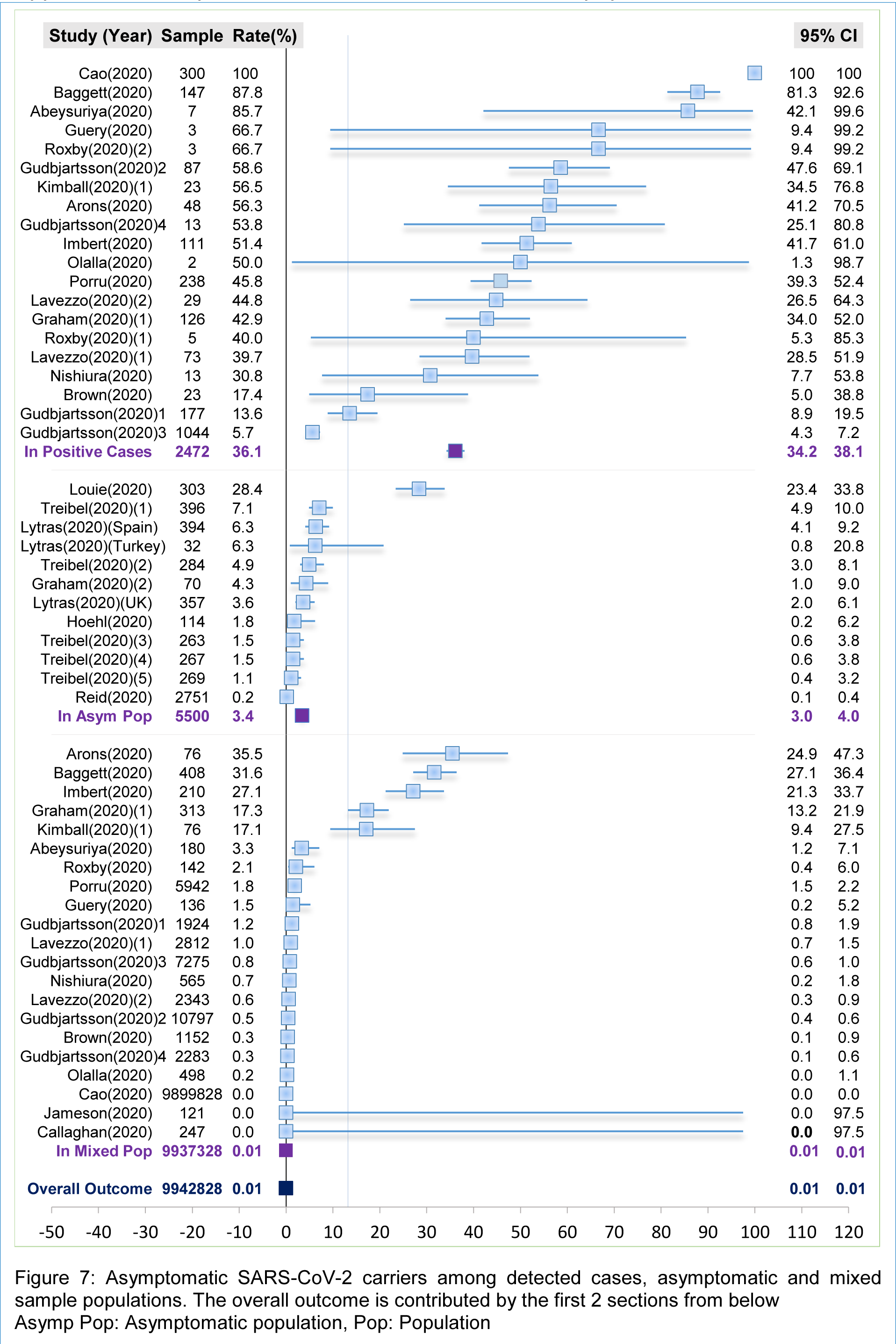
Graph of Outcome Behaviour in different populations

