## Supplemental Material 1 for "Mass Testing with Contact Tracing Compared to Test and Trace for Effective Suppression of COVID-19 in the UK: A rapid review"

### Synthesis Without Meta-analysis (SWiM) reporting items

The citation for the Synthesis Without Meta-analysis explanation and elaboration article is: Campbell M, McKenzie JE, Sowden A, Katikireddi SV, Brennan SE, Ellis S, Hartmann-Boyce J, Ryan R, Shepperd S, Thomas J, Welch V, Thomson H. Synthesis without meta-analysis (SWiM) in systematic reviews: reporting guideline BMJ 2020;368:l6890 http://dx.doi.org/10.1136/bmj.l6890

| **SWiM is intended to complement and be used as an extension to PRISMA** | | | |
|---|---|---|---|
| **SWiM reporting item** | **Item description** | **Page in manuscript where item is reported** | **Other*** |
| *Methods* | | | |
| **1** Grouping studies for synthesis | 1a) Provide a description of, and rationale for, the groups used in the synthesis (e.g., groupings of populations, interventions, outcomes, study design) | 10 | |
| | 1b) Detail and provide rationale for any changes made subsequent to the protocol in the groups used in the synthesis | N/A | |
| **2** Describe the standardised metric and transformation methods used | Describe the standardised metric for each outcome. Explain why the metric(s) was chosen, and describe any methods used to transform the intervention effects, as reported in the study, to the standardised metric, citing any methodological guidance consulted | 10, 27 | |
| **3** Describe the synthesis methods | Describe and justify the methods used to synthesise the effects for each outcome when it was not possible to undertake a meta-analysis of effect estimates | 10, 27 | |
| **4** Criteria used to prioritise results for summary and synthesis | Where applicable, provide the criteria used, with supporting justification, to select the particular studies, or a particular study, for the main synthesis or to draw conclusions from the synthesis (e.g., based on study design, risk of bias assessments, directness in relation to the review question) | 11, 29 | |

#### Synthesis Without Meta-analysis (SWiM) reporting items

| SWiM reporting item | Item description | Page in manuscript where item is reported | Other* |
|---|---|---|---|
| **5** Investigation of heterogeneity in reported effects | State the method(s) used to examine heterogeneity in reported effects when it was not possible to undertake a meta-analysis of effect estimates and its extensions to investigate heterogeneity | 10, 34 | |
| **6** Certainty of evidence | Describe the methods used to assess certainty of the synthesis findings | 11, 28, 30 | |
| **7** Data presentation methods | Describe the graphical and tabular methods used to present the effects (e.g., tables, forest plots, harvest plots).<br><br>Specify key study characteristics (e.g., study design, risk of bias) used to order the studies, in the text and any tables or graphs, clearly referencing the studies included | 11, 13 to 26 | |
| *Results* | | | |
| **8** Reporting results | For each comparison and outcome, provide a description of the synthesised findings, and the certainty of the findings. Describe the result in language that is consistent with the question the synthesis addresses, and indicate which studies contribute to the synthesis | 27 to 33, | |
| *Discussion* | | | |
| **9** Limitations of the synthesis | Report the limitations of the synthesis methods used and/or the groupings used in the synthesis, and how these affect the conclusions that can be drawn in relation to the original review question | 35 | |

PRISMA=Preferred Reporting Items for Systematic Reviews and Meta-Analyses.
*If the information is not provided in the systematic review, give details of where this information is available (e.g., protocol, other published papers (provide citation details), or website (provide the URL)).
